# A genome-wide pleiotropy study between atopic dermatitis and neuropsychiatric disorders

**DOI:** 10.1101/2024.10.30.24316209

**Authors:** Charalabos Antonatos, Alexandros Pontikas, Adam Akritidis, Dimitra Mitsoudi, Sophia Georgiou, Alexander J. Stratigos, Aikaterini Zacharopoulou, Stamatis Gregoriou, Katerina Grafanaki, Yiannis Vasilopoulos

## Abstract

Comorbidities between atopic dermatitis (AD) and neuropsychiatric disorders are frequently reported, however the extent of shared genetic architecture remains unclear. Here, we performed a large-scale genome-wide pleiotropy approach to investigate the genetic correlations and causal associations between AD and five neuropsychiatric disorders, attention deficit hyperactivity disorder (ADHD), autism spectrum disorder (ASD), bipolar disorder (BP), major depressive disorder (MDD), and schizophrenia (SCZ). Using genome-wide association (GWAS) data, we explored genetic overlaps, pleiotropic loci and assessed the capacity of pleiotropic associations to identify drug targets. We identified significant positive genetic correlations between AD and ADHD (rg=0.14, P-value=2×10), MDD (rg=0.13, P-value=1.2×10 ³) and BP (rg=0.11, P-value= 4×10 ³). Genome-wide pleiotropy scans identified 37 distinct pleiotropic loci between AD and neuropsychiatric traits, with gene-based analyses highlighting 86 unique genes participating in inflammatory pathways. Pleiotropy-informed target prioritization facilitated the identification of novel pathophysiological mechanisms for AD and putative drug targets, such as members of TNF and JAK-STAT3 signaling. Mendelian randomization provided evidence of a causal relationship between genetic liability to MDD and BP with an increased risk of AD, independent of sample overlap. Collectively, our findings elucidate shared molecular mechanisms between AD and neuropsychiatric disorders, emphasizing immune-related pathways as key contributors to both disease categories, with potential implications for therapeutic interventions targeting common inflammatory mechanisms.

## Introduction

Atopic dermatitis (AD) is a common, inflammatory skin disease affecting over 2% of the global population, with rising lifetime prevalence in developed countries[1,2]. AD is primarily characterized by intense pruritus and inflammatory cutaneous lesions, often exacerbated by bacterial colonization such as *Staphylococcus aureus*, further aggravating the disease and diminishing patients’ quality of life[2]. A key pathogenic feature of AD is the disruption of the epidermal barrier linked to null *FLG* mutations that increase skin permeability, and a concomitant type 2 immune response driven by elevated allergen penetrance. Recent genome-wide association studies (GWASs) have elucidated a proportion of the disease heritability, estimated at 80%, uncovering 81 independent risk loci in over 800000 participants of European descent[3]. Some of these loci are associated with both atopic phenotypes[4] and mental health[5].

Multiple studies have linked AD with neuropsychiatric disorders. For instance, our recent findings suggest that genetic liability to feeling worried may increase risk of AD[6]. Mechanistically, this is hypothesized to involve the skin-brain axis, where psychological stress[7] impacts AD via neuroinflammation. Type 2 cytokines, in particular, promote sensory nerve growth and thus sustain the itch-scratch cycle[8, 9]. Patients with AD frequently experience elevated rates of sleep disturbances, including short sleep duration and early awakenings[10]. Intense pruritus and social stigmatization further contribute to neuropsychiatric outcomes. Children with severe AD have a two-fold risk of developing depressive symptoms independently of sleep quality and inflammatory biomarkers[11], while mild-to-moderate AD has been associated with increased prevalence of attention deficit hyperactivity disorder (ADHD) and bipolar disorder (BP)[12]. Contrastingly, a population-based cohort study of more than 400000 children with AD found null associations between AD and neuropsychiatric disorders[13]. Similarly, a mendelian randomization (MR) study revealed no bidirectional relationships between AD and major depressive disorder (MDD)[14]. Understanding the extent to which AD and neuropsychiatric disorders share similar genetic components may facilitate the interpretation of the above contradictory signals. Cross-trait methodological approaches, as performed in several genetic studies, offer the potential to dissect shared genetic and molecular mechanisms[15], provide novel insights in disease pathophysiology[16], while pleiotropic loci may serve as novel interventional targets[17].

In the current study, we used large-scale GWASs to evaluate the pleiotropic associations between AD[3] and five discrete neuropsychological outcomes, ADHD[18], autism spectrum disorder (ASD)[19], BP[20], MDD[21] and schizophrenia (SCZ)[22]. We examined the impact of single nucleotide polymorphisms (SNPs) in common genetic associations between AD and neuropsychiatric disorders through genome-wide pleiotropy scans and explored shared mechanisms at the gene and pathway levels. Using a recently developed pleiotropy-informing algorithm, we evaluated whether pleiotropic associations between AD and neuropsychiatric disorders could elucidate pathway crosstalks with implications in therapeutic development. Finally, we examined the pairwise causal relationships in the skin-brain axis of AD through MR. Ultimately, our findings provide valuable biological insights in the complex relationship between neuropsychiatric disorders and AD, underscoring potential links through inflammatory mechanisms within the skin-brain axis.

## Methods

### Study design

We utilized GWAS summary statistics for AD, ADHD, ASD, BP, MDD and SCZ. For each trait, we selected the largest GWAS conducted in participants of European ancestry. Details for each summary statistics used in the study is provided at Table S1. Ethical approval and informed consent from participants are documented in the respective GWAS. Data collection spanned from November 2023 to January 2024. All analyses were performed using the hg19 (GRCh37) genome assembly.

### Statistical analyses

All analyses began in February 2024. Details regarding all methods can be found at Supplementary File. To begin, we estimated global genetic correlation patterns between AD and each neuropsychiatric disorder using linkage disequilibrium score regression (LDSC)[23]. Statistical significance was defined at a P-value threshold of 0.01 after applying Bonferroni correction for multiple comparisons.

We restricted analyses in common, biallelic variants (minor allele frequency (MAF)>0.01) outside the major histocompatibility complex (MHC) region to avoid complex LD patterns. Due to the presence of sample overlap in some GWAS summary statistics, we applied appropriate methodological corrections to mitigate potential bias. Specifically, we used PLACO to identify pleiotropic associations between AD and each neuropsychological disorder[24]. Variants with Z-score^2^<80 were excluded to avoid spurious results as recommended by the authors. Prior to employing PLACO, we de-correlated the Z-scores due to potential bias inserted by sample overlap[24]. Significant pleiotropic variants were defined by a P-value threshold of <5×10^-8^.

Genome-wide pleiotropy results were next submitted to the functional mapping and annotation of genetic associations (FUMA) platform[25]. FUMA was used to (i) identify independent pleiotropic loci based on a two-step clumping approach (Supplementary File), and (ii) perform functional annotation of pleiotropic variants with ANNOVAR[26]. In each pleiotropic locus, we further identified a credible set of variants that were 99% likely to contain causal variants through FM-summary[27]. Pairwise colocalization analysis was performed using coloc-v5.2.3 on the independent pleiotropic loci between AD and each neuropsychiatric disorder[28]. We analyzed ±200 kb regions around lead SNPs, declaring loci with posterior probability (PP)H4>0.7 as colocalized. When examining all non-overlapping pleiotropic loci between AD and neuropsychiatric traits, loci were defined through the intersection of each pairwise pleiotropic locus. The SNP with the highest PP.H4 was considered the most likely causal variant.

The PLACO results were further submitted to gene-level and gene-set analyses with MAGMA v1.10 software[29]. The summation of sample sizes in each pairwise trait was used as an input for MAGMA v1.10. For MAGMA gene-level analysis, we used predefined gene boundaries based on the Ensembl v110 platform[30], while gene-set analysis was conducted with the molecular signatures database (MSigDB) v2023.1 human collections[31].

We next implemented a recently developed algorithm that leverages pleiotropic associations between traits to evaluate whether pleiotropic variants could aid early-stage drug development and repurposing. The pleiotropy-informing prioritization and evaluation (PIPE) employs genomic proximity, chromatin conformation and gene expression to prioritize candidate target genes at the pleiotropic loci[17]. In total, we applied PIPE to all 5 pairwise comparisons and evaluated its performance in a three-step process. PIPE integrates novel metrics to assess whether a prioritized gene list can be used to develop therapeutic targets, named pleiotropy-informing clinical therapeutics (PICT). This metric is derived by the leading prioritization analysis (LPA), a methodology analogous to gene set enrichment analysis evaluating the capacity of a gene list to be a clinical proof-of-concept target (Supplementary File). Next, we explored the skin-brain axis in AD by merging all 5 pairwise comparisons and performing functional enrichment with Kyoto Encyclopedia of Genes and Genomes (KEGG) pathways database[32]. We also examined the potential druggability of prioritized genes through constructing a supra-hexagonal map[33]. Genes were classified as targetable if a known protein structure, derived from the protein databank (PDB) database[34], was predicted to contain druggable pockets through the fpocket software[35]. Independent SNPs from the FUMA algorithm were used as PIPE input for each pairwise comparison.

Finally, we conducted bidirectional, univariable MR analyses to investigate potential causal trait relationships of AD with neuropsychiatric traits. The major analysis outcome was declared as the inverse-variance weighted (IVW) method[36], while sensitivity analyses were employed to evaluate the robustness of the results including MR-Egger, weighted median and MR-PRESSO[6]. A Bonferroni-corrected P-value of 0.01 was set as the significance threshold. In cases of sample overlap and nominally significant results (P-value<0.05), we employed MRlap, a recently developed tool to account for potential sample overlap in univariable MR studies. For MRlap, we excluded variants identified as outliers from MR-PRESSO.

## Results

### Genetic correlations between atopic dermatitis and neuropsychiatric disorders

The strongest genetic correlation was observed between AD and ADHD (r_g_=0.14, P-value=2.57×10^-5^), followed by MDD (r_g_=0.13, P-value=3.3×10^-6^) and BP (r_g_=0.11, P-value=1.8×10^-4^). Genetic correlations between AD and SCZ (r_g_=0.05, P-value=1.89×10^-2^) and ASD (r_g_=0.13, P-value=2.31×10^-2^) achieved suggestive significance (Table S2).

### Pleiotropic loci between atopic dermatitis and neuropsychiatric disorders

In total, 53 pleiotropic loci were identified between all neuropsychiatric disorders and AD (Fig. 1; Table S3), reduced to 37 non-overlapping. Quantile-quantile plots are shown in Fig. S1. The largest number of pleiotropic loci was identified between AD and SCZ (n=15), while the lowest number was observed between AD and ASD (n=5) (Table S3). As expected, some pleiotropic loci were shared in >1 pairwise trait, such as the *TRAF3* locus shared among ADHD, BP and MDD (Fig. 1). The modest genetic correlation observed earlier was further validated in pleiotropy analyses. Specifically, 5/11 loci (45.45%) showed concordant effects in the AD-ADHD pair, 3/5 (60%) in the AD-ASD pair, 7/11 (63.63%) in the AD-BP pair, 6/11 (54.54%) in the AD-MDD pair and 7/15 (46.66%) in the AD-SCZ pair (Table S3). Functional enrichment analysis with ANNOVAR indicated that most variants were mapped in intronic regions (Table S4), where the maximum proportion was observed in the AD-BP trait pair (71.8%).

**Fig. 1.**
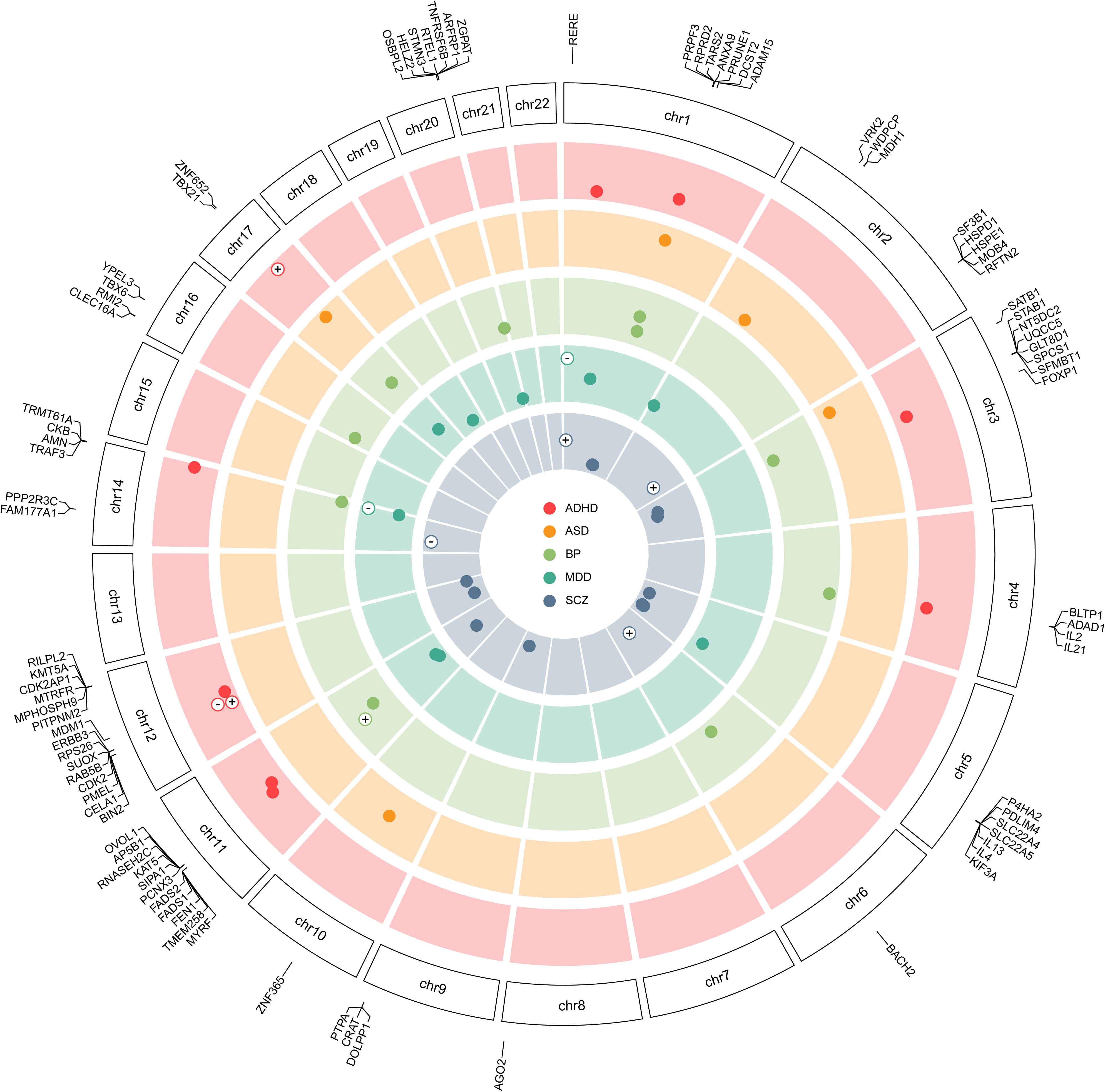
Circular plot of pleiotropic loci between atopic dermatitis and neuropsychiatric disorders. Each layer corresponds to independent risk loci identified from the FUMA pipeline (Table S3). Nodes colored in white show risk loci with a posterior probability of H4 (PP.H4) > 0.7 (Table S5). The outer layer corresponds to genes derived from the MAGMA gene-level analysis (Table S8). A plus sign (+) is assigned to variants with concordant sign in AD and neuropsychiatric disorder, while a minus sign (-) is assigned to variants with discordant sign (Table S3). ADHD, attention-deficit hyperactivity disorder; ASD, autism spectrum disorder; BP, bipolar disorder; MDD, major depressive disorder; SCZ, schizophrenia.

Through fine-mapping, we identified several candidate causal SNPs for pleiotropic loci between AD and each neuropsychiatric disorder (Table S5). Colocalization analyses provided additional support for several pleiotropic associations between AD and neuropsychiatric diseases (Fig. 1; Table S6). Pairwise colocalizations indicated the presence of pleiotropic associations at *RERE* locus in both AD-MDD and AD-SCZ pairs at a posterior probability H4 (PP.H4)>0.7, a previously identified risk locus for all 3 diseases[3, 21, 22](Fig. 1). This locus was further validated when examining all 37 non-overlapping risk loci (Table S7). Locus compare and locus zoom plots are available at Fig. S2.

### Prioritization of candidate pleiotropic genes and pathways

Using MAGMA v1.10 software, we identified 116 (86 unique) genes mapped in pleiotropic loci across all trait pairs at a pairwise-specific Bonferroni-corrected P-value threshold (Table S8). Similarly, several genes were shared among >2 trait pairs (Fig. 1). For example, *TRAF3* and *IL4* were shared among 4/5 trait pairs excluding AD-ASD, while *TNFRSF6B* and *ARFRP1* and *AMN* were shared among neuropsychiatric disorders excluding AD-SCZ and AD-ADHD pairwise comparisons (Table S8). Gene-set analysis using the MSIGDB v2023.1HS reported several shared inflammatory pathways between potential pleiotropic gene lists (Table S9). For instance, interleukin (IL)21 production was shared among AD-ADHD and AD-SCZ pairs, while leukocyte differentiation was enriched in the AD-ADHD, AD-BP and AD-MDD pairs (Table S9).

### Pleiotropy-informed therapeutic prioritization

Prioritized gene lists from all trait pairs can be found at Table S10. To evaluate the performance of PIPE in each pairwise comparison, we adopted a three-step sensitivity analysis. First, we examined the ability of PIPE-derived prioritized genes in identifying novel therapeutic targets. In all comparisons, PIPE reported an increased discriminative ability on identifying clinical proof-of-concept targets against simulated negative targets (PIPE0), as evidence by higher area under the curve (AUC) scores, with the largest difference observed in the AD-SCZ pair (Table S11). Next, we evaluated the capacity of prioritized gene lists as proof-of-concept therapeutic targets through the LPA and quantified by the PICT metric across all pairwise comparisons. We observed significant enrichment scores at a false discovery rate (FDR) threshold of 0.05 in all pairwise comparisons, while the largest PICT score was obtained in the AD-SCZ pair (Table S12). Finally, we examined the relationships between each pairwise comparison at the gene level. Member genes for each disease were defined as the clinical proof-of-concept targets recovered at the LPA (Table S15). Several genes reported overlaps in trait pairs, including *NR3C1* in AD-BP, AD-MDD and AD-SCZ pairs, *MTNR1A* in AD-ASD, AD-MDD and AD-SCZ pairs and *VDR* in AD-ASD and AD-SCZ pairs (Fig. 2a; Table S12).

**Fig. 2.**
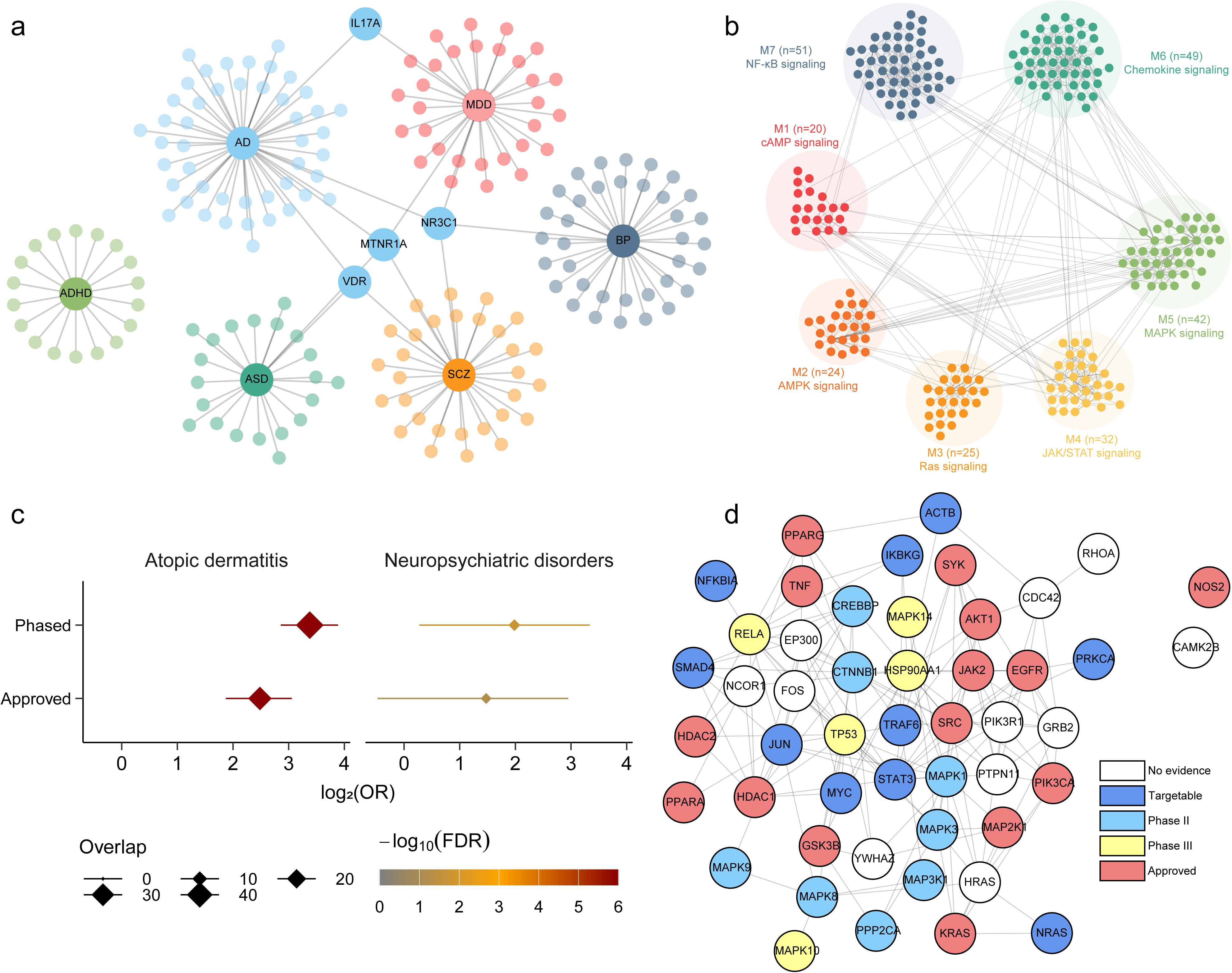
Modular analysis of pathway crosstalks and prioritization map in atopic dermatitis and neuropsychiatric disorders. (a) Identification of target genes at the pathway crosstalk level between AD and neuropsychiatric disorders. Genes linked to >1 disease are shown in black font (Table S12). (b) Visualization of modular analysis (Table S13) and results of pathway enrichment analysis (Table S14). (c) Forest plot indicating the one-sided Fisher’s exact test enrichment for approved drug targets in Modules 3-7. (d) Protein-protein interaction network of the C2 cluster in STRING v12.0 database (Table S17). Color of each node represents the clinical phase of drugs targeting each gene, as derived from the CheMBL database. ADHD, attention-deficit hyperactivity disorder; ASD, autism spectrum disorder; BP, bipolar disorder; MDD, major depressive disorder; SCZ, schizophrenia; OR, odds ratio; FDR, false discovery rate.

For each pairwise comparison, we derived pathway crosstalks in each prioritized gene list by integrating credit score information with pathway-based gene-gene interactions from the KEGG pathways (Fig. S3-S7). We next merged all pathway crosstalks to identify prioritized modules associated with the skin-brain axis in AD. We identified 7 discrete modules (Fig. 2b; Table S13) with a spin-glass model and simulated annealing and performed a pathway enrichment analysis with KEGG pathways. The modules M3-M7 were associated with discrete inflammatory pathways interplaying in AD pathogenesis, while Modules M1-M2 were enriched in neuropsychiatric diseases-related pathways such as cAMP and AMPK signaling (Fig. 2b; Table S14). Disease enrichment analysis with the DisGeNET[38] platform further linked M3-M7 modules with AD (Table S15). Notably, significant enrichment for approved and phased drugs in AD was observed in M3-M7, as derived from the ChEMBL database, contrary to all neuropsychiatric disorders with 95% confidence intervals (CIs) overlapping the null (Fig. 2c; Table S16).

Finally, to assess potentially novel drug candidates for AD, we constructed a prioritization map based on the pathway crosstalk of all pairwise comparison. Genes were clustered with the supraHex package in 3 different clusters (C1-C3; Table S17). We hypothesized that clusters reporting a consistently higher credit score in all pairwise comparisons would be more relevant for AD, contrary to pair-specific scores. Cluster C2 displayed stronger associations across all pairwise comparisons (Fig. S8) as well as the highest proportion of targetable genes (89.8%; Figs. S9). Moreover, genes in the C2 cluster reported significant direct protein-protein interactions through the STRING v12.0 database (Fig. 2d). We observed several drug targets previously associated with the pathogenesis of AD, such as TNF and JAK2, while genes relevant to AD were also identified as structurally targetable, including STAT3 and PPARA (Fig. 2d). Structurally targetable domains of each targetable protein are provided at Fig. S10.

### Bidirectional mendelian randomization

Bidirectional MR analyses were employed to evaluate pairwise causal associations between AD and neuropsychiatric diseases. Instrumental variables (IVs), results and sensitivity analyses are provided in Tables S18-S23. Although genetic liability to AD was not associated with any neuropsychiatric disease (Table S19; Fig. S11), genetic liability to BP was associated with increased AD risk (OR_IVW_ (95% CI)=1.06 (1.01-1.08); Fig. S12) with consistent estimates in all sensitivity analyses (Fig. S12). MR-PRESSO further indicated that genetic liability to MDD was associated with increased AD risk (OR_MR-PRESSO_ (95% CI)=1.14 (1.04-1.25); Fig. S12). Since BP, MDD and AD consisted of UKB samples, we applied MRlap, a recently developed tool to correct for potential sample overlap in MR analyses excluding outliers detected from MR-PRESSO. Strikingly, sample overlap underestimated the effect estimates in each case (Table S23). In specific, genetic liability to MDD displayed a significant association with AD OR_MRlap_ (95%CI)=1.07 (1.02-1.12)), while genetic liability to BP retained causal estimates to AD after correction to sample overlap (OR_MRlap_ (95%CI)=1.06 (1.02-1.11)).

## Discussion

In this work, we systematically interrogated genome-wide summary data to understand the shared genetic basis underlying AD and neuropsychiatric disorders. We identified significant, albeit modest global genetic correlation between AD and 3/5 neuropsychiatric disorders. By leveraging pleiotropic associations and molecular evidence, we uncovered novel mechanisms that participate in the skin-brain axis suggesting new therapeutic targets. Finally, we establish a putative causal role of MDD and BP on AD risk.

The epidemiological evidence linking neuropsychiatric disorders with AD is well established, with AD patients frequently displaying higher levels of anxiety and depressive symptoms[11,12,39,40]. Depression, the most extensively studied psychiatric comorbidity in AD, is hypothesized to result from both social stigmatization experienced by AD patients and intersection of inflammatory pathways[5], while fewer studies examining the reverse association[41]. Our findings support a causal effect of genetic liability to MDD on AD risk, a result robust across sensitivity analyses and after adjusting for sample overlap[37]. The link between AD and severe mental illnesses, such as BP, remains less well-defined[41]. A single study has reported a higher prevalence of BP in AD patients[39], however the association was attenuated when adjusting for possible confounders such as glucocorticoid use in an independent study[43]. In our analysis, genetic liability to BP was causally associated with AD, which held across all sensitivity analyses. However, recent MR approaches have contradicted observational findings, displaying null bidirectional relationships of AD with MDD[14, 44]. Moreover, Cao and colleagues further reported nominally significant causal effect of AD on ASD (P-value_IVW_=0.026) and ADHD (P-value_IVW_=0.033) risk[44], findings we did not replicate (Table S19), although we detected a significant MR-Egger intercept in the AD-ADS comparison (Table S22). In contrast to prior studies, we used (i) a larger number of IVs, (ii) summary statistics from larger GWAS meta-analyses, as well as (iii) correction for potential bias through sample overlap. Despite this, effect sizes were modest (Table S19), implying low statistical power to detect such associations. The above discrepancies stress the need for investigating disease endotypes and stratification by disease severity. Severe AD may be exacerbated by poor mental health outcomes[11, 45], while milder AD was associated with ADHD and ASD, a discrepancy potentially explained by the administration of systemic treatments in severe cases ameliorating neuropsychiatric comorbidities[12].

The positive r_g_ between AD and 3/5 neuropsychiatric disorders (Table S2) prompted us to dissect their shared genetic etiology. We identified 37 non-overlapping pleiotropic loci between AD and all 5 neuropsychiatric disorders, distributed across the genome, with some loci shared in >2 pairwise comparisons (Fig. 1; Table S3). Genes such as *TRAF3* and *IL4* were shared among 4/5 comparisons excluding ASD (Table S8). *TRAF3*, a previously established AD risk gene[46], holds a versatile activity in immune-related functions, particularly within NF-kB signaling[47]. Variants in *TRAF3* have been also linked to neuropsychiatric disorders through multi-trait GWAS analysis[48]. On the contrary, *IL4* facilitates T helper (T_H_)2 polarization[49] and IgE production[50], hence serving as an important regulator of AD pathogenesis. Patients treated with dupilumab, a monoclonal antibody blocking IL4RA, have reported significant improvements in mental health outcomes[51] such as sleep disturbances[52], anxiety and depressive symptoms[53], as well as reduced psychostimulant use in AD-ADHD comorbidity[54]. A similar pattern was observed in gene-set analyses, where inflammatory pathways governed the pleiotropic evidence between AD and each neuropsychiatric trait (Table S9). Examples such as IL21 signaling in AD-ADHD and AD-SCZ pairs, as well as leukocyte differentiation in AD-ADHD, AD-BP and AD-MDD pairs suggest shared potential inflammatory mechanisms at both the gene and pathway level.

By leveraging pairwise pleiotropic associations, we constructed an inflammatory-driven network underlying the comorbidity between AD and neuropsychiatric disorders (Fig. 2). All pairwise prioritized gene lists were strong predictors for clinical proof-of-concept targets in AD (Table S11). We highlight some potential members for experimental validation and follow-up studies. For instance, MAPK signaling has been associated with *FLG* and *IVL* expression through IL17, a gene emerged as a potential mediator between AD and MDD (Fig. 2a)[55]. Tumor necrosis factor (TNF) alone can induce AD-like cutaneous symptoms including skin permeability and increased free fatty acid (FFA) concentrations[56], while hippocampal TNF signaling is required to preserve anxiety behaviors[57]. In experimental models, blockage of TNF-like weak inducer of apoptosis (TWEAK) reported similar amelioration of AD symptoms and transcriptomic signals compared to IL13 inhibition[58], hence suggesting the potential utility of TNF signaling members in AD drug administration. Using supra-hexagonal clustering (Fig. 2d), we identified additional candidate genes in the C2 cluster, participating in AD pathogenesis and possibly in the skin-brain axis. Members of TNF and NF-kB signaling pathways with druggable structures were identified including *RELA*, *NFKBIA*[59] and *TRAF6*[60], along with *JAK2*, an approved drug target for AD [61](Fig. 2d). Of particular interest is *STAT3*, a transcription factor that directly interacts with TRAF6 and JAK2 (Fig. 2d) and holds a tissue-specific role in AD pathogenesis. Conditional ablation of *STAT3* in mice leads to AD-like cutaneous inflammation and TSLP upregulation in keratinocytes[62], and modulates KLK5 and SPINK5 keratinocyte expression involved in epidermal barrier integrity[63]. Conversely, STAT3 activation in sensory nerves promotes inflammatory itch[64], while astrocytic STAT3 inhibition alleviates chronic itch in AD-like mice[65]. Thus, STAT3 exhibits a dual role in AD pathogenesis, with the JAK-STAT3 axis serving as a potential drug target[66, 67].

Our study has caveats. First, all GWAS data used were restricted to participants of European ancestry, thereby limiting the generalizability to other ethnic groups. Second, rare variants (MAF < 1%) and variants within the MHC were excluded, potentially overlooking pleiotropic associations. Third, sample overlap in GWAS may have introduced bias in our results. To partially tackle this, we applied appropriate methodological approaches to mitigate sample overlap bias, including de-correlation of Z-scores prior to PLACO and MRlap as a sensitivity analysis. Fourth, we examined statistical pleiotropy, limiting our ability to distinguish between true pleiotropy (that is, a variant is associated with 2 traits), and biological pleiotropy (that is, a gene is associated with 2 traits through different biological pathways)[68]. Fifth, the PIPE algorithm incorporates molecular evidence from immune and brain-related tissues. Despite the apparent application of these evidence in AD, we cannot rule out the contribution of skin-specific effects in the relationship between AD and neuropsychiatric disorders. Nonetheless, several pathways highlighted in our study have an established role in AD pathogenesis through keratinocytes, including JAK-STAT, TNF and MAPK signaling.

In conclusion, we elucidate the shared genetic architecture between AD and neuropsychiatric disorders. Some of these shared loci, such as *TRAF3* and *IL4*, support a role for inflammatory pathways governing comorbidity estimates. Our pleiotropy-informed approach to therapeutic target identification has revealed novel pathophysiological mechanisms and putative drug targets for AD. By leveraging genetic and molecular data, our study advances the understanding of skin-brain axis underlying neuropsychiatric comorbidities in AD.

## Acknowledgements

CA was financially funded by the “Andreas Mentzelopoulos Foundation”.

## Funding

This investigation was supported in part by a research grant from the National Eczema Association NEA23-CRG186.

## Author Contributions

Conceptualization: CA, StG, KG; Methodology: CA, DM; Investigation: CA, AP, AA, DM; Visualization: CA; Supervision: YV, StA: Writing-Original Draft: CA; Writing-review and editing: CA, AP, AA, DM, SG, AJS, AZ, StG, KG, YV

## Competing interests

The authors have no conflict of interest to declare.

## Data and materials availability

Summary statistics can be downloaded from the GWAS catalog (https://www.ebi.ac.uk/gwas/) and the Psychiatric Genomics Consortium (PGC; https://pgc.unc.edu/). Summary statistics from each pleiotropic pairwise comparison will be made available during peer review, and publicly available after publication of the manuscript.

## Supplementary Figures

Fig. S1. Quantile-Quantile (QQ) plots of all pairwise comparisons.

Fig. S2. Locus zoom and locus compare plots for all significant colocalizations in pairwise comparisons

Fig. S3. Pathway crosstalk visualization based on pleiotropic evidence in AD-ADHD pairwise comparison. Parentheses in each node represent the rank in the prioritized gene list.

Fig. S4. Pathway crosstalk visualization based on pleiotropic evidence in AD-ASD pairwise comparison. Parentheses in each node represent the rank in the prioritized gene list.

Fig. S5. Pathway crosstalk visualization based on pleiotropic evidence in AD-BP pairwise comparison. Parentheses in each node represent the rank in the prioritized gene list.

Fig. S6. Pathway crosstalk visualization based on pleiotropic evidence in AD-MDD pairwise comparison. Parentheses in each node represent the rank in the prioritized gene list.

Fig. S7. Pathway crosstalk visualization based on pleiotropic evidence in AD-SCZ pairwise comparison. Parentheses in each node represent the rank in the prioritized gene list.

Fig. S8. Pathway crosstalk prioritization map. (a) Illustration of the supra-hexagonal map with mapped all 3 identified clusters. Each cluster is separated by a space. (b) Prioritization map colored by credit score in each pairwise comparison. ADHD, attention deficit hyperactivity disorder; ASD, autism spectrum disorder; BP, bipolar disorder; MDD, major depressive disorder; SCZ, schizophrenia.

Fig. S9. Proportion of structurally targetable genes in each supra-hexagonal cluster (C1-C3) as identified with the fpocket software.

Fig. S10. Structurally targetable proteins in the C2 cluster. The x axis represents structurally targetable domains of each protein with known protein structure through the PDB database in the y axis.

Fig. S11. Mendelian randomization estimates when atopic dermatitis was set as exposure. Fig. S12. Mendelian randomization estimates when atopic dermatitis was set as outcome.

## Supplementary Tables

Table S1. Genome-wide summary statistics used in the study

Table S2. Global genetic correlations between atopic dermatitis and mental diseases.

Table S3. Independent pleiotropic loci between atopic dermatitis and mental diseases.

Table S4. ANNOVAR enrichment analysis results for each pairwise comparison.

Table S5. Cumulative summation of probability of SNP in the 99% credible set from fine-mapping in each pleiotropic association for pairwise comparisons.

Table S6. Colocalization results for each pairwise comparison in pairwise-specific pleiotropic loci.

Table S7. Colocalization results for each pairwise comparison in all non-overlapping pleiotropic loci.

Table S8. MAGMA gene-level analyses results for each pairwise comparison.

Table S9. MAGMA gene-set analyses results for each pairwise comparison.

Table S10. Credit scores in all pairwise comparisons.

Table S11. Performance of PIPE in identifying clinical proof-of-concept targets from simulated negative targets.

Table S12. Leading priotization analysis and PICT scores for each pairwise comparison.

Table S13. Pathway crosstalk gene members and functional modules.

Table S14. Significant pathway enrichment analysis results for each pathway crosstalk gene module

Table S15. Significant disease enrichment analysis results for each pathway crosstalk gene module

Table S16. One-tailed Fisher’s exact test of approved or phased drug target enrichment in M3-M7.

Table S17. Pathway crosstalk-based prioritization map

Table S18. Summary statistics of the instrumental variables used in our analyses. Table S19. Results of all analyses conducted.

Table S20. Heterogeneity and I2GX metrics for all analyses. Table S21. Outliers detected from MR-PRESSO.

Table S22. Results of the MR-Egger intercept.

Table S23. Correction for sample overlap with MRlap.

## Notes

### Competing Interest Statement

The authors have declared no competing interest.

### Author Declarations

GWAS summary statistics are publicly available from the GWAS catalog (https://www.ebi.ac.uk/gwas/home) and the PGC consortium (https://pgc.unc.edu/).

