## Supplementary Methods for "A genome-wide pleiotropy study between atopic dermatitis and neuropsychiatric disorders"

**Charalabos Antonatos *et al*.**

**Supplementary methods**

**Data resources**

We leveraged large-scale GWASs of European participants for atopic dermatitis (AD)[1], attention deficit hyperactivity disorder (ADHD)[2], autism spectrum disorder (ASD)[3], bipolar disorder (BP)[4], major depressive disorders (MDD)[5] and schizophrenia (SCZ)[6]. Ethical approval was obtained in all original studies. Sample collection, quality control and imputation methodology for each study is available in the corresponding publications. Since several of the datasets display sample overlap, primarily due to the inclusion of UK biobank (UKB) data, all methods handled appropriately sample overlap.

**Genome-wide genetic correlations**

We calculated SNP-based bivariate genetic correlations (*r_g_*) between AD and neuropsychiatric disorders using linkage disequilibrium (LD) score regression (LDSC) which produces *r_g_* estimates unbiased by sample overlap[7]. We used precomputed LD score estimates derived from the European 1000 Genomes project, calculated in a HapMap 3 SNP panel of ~1.2 million common variants. Multiple testing was adjusted using the Bonferroni correction method (P<0.05/5=0.01).

**Pleiotropy analysis under the composite null hypothesis**

For each neuropsychiatric disorders and AD, we employed the pleiotropy analysis under the composite null hypothesis (PLACO)[8]. PLACO assesses pleiotropy at the single-SNP level, partitioning the composite null hypothesis into three distinct sub-null hypotheses: (i) H_00_, where the variant is not associated with any trait, (ii) H_10_, where the variant is associated only with the first trait, and (iii) H_01_, where the variant is associated only with the second trait. The alternative hypothesis H_11_ assumes that the variant is associated with both traits. For PLACO, we included only (i) shared, bi-allelic variants (ii) with minor allele frequency (MAF) larger than 0.01 and (iii) outside the extended major histocompatibility complex (MHC) (chr6:25000000-33000000) due to complex linkage disequilibrium (LD) patterns. Variants were harmonized between datasets with the bigsnpr v1.12.4 R package to ensure the robust assessment of pleiotropy, while variants displaying Z-score^2^>80 were further excluded to avoid spurious pleiotropic signals. To alleviate potential bias due to sample overlap, we first de-correlated Z-scores prior to applying the pleiotropy analysis. To estimate correlation patterns between Z-scores, PLACO borrows variants with P-value<10^-4^ and calculates the Pearson correlation parameter *ρ*. SNPs with P_PLACO_<5×10^-8^ were considered as statistically significant pleiotropic variants.

**Functional characterization of pleiotropic loci**

We adopted a two-stage clumping approach for the identification of pleiotropic loci. Genome-wide significant (GWS) SNPs were first clumped at an r^2^ threshold < 0.6 to identify independent significant variants. The ANNOtate VARiation (ANNOVAR)[9] software was used to map independent significant variants to the nearest protein-coding gene (Ensembl build v110) at a maximum distance of 10kb. Pleiotropic risk loci were defined after assigning independent significant SNPs in LD at 0.1≤ r^2^≤ 0.6. If independent significant SNPs were closer than 250kb, the pleiotropic risk loci were merged in one locus. Lead SNPs were identified through a second clumping of the independent significant variants at r^2^ < 0.1. The European sample of 1000 Genomes Project Phase 3] was used as a reference panel to calculate pairwise LD between SNPs using PLINK v1.9. Functional analyses were conducted with the functional analysis and annotation of GWASs (FUMA) platform, v1.6.1[10].

**Fine-mapping and Bayesian colocalization analysis**

The lead variant does not necessarily refer to the causal variant in each locus. Therefore, we applied FM-summary, a fine mapping method that requires only summary statistics[11]. FM-summary identifies a credible set of SNPs at a 99% posterior probability to contain causal variants at each predefined pleiotropic locus. FM-summary focuses on mapping the primary signal (e.g., SNP with the lowest P-value) and applies a flat prior distribution with a steepest descent approximation, under the assumption of at least one causal variant in each region.

We implemented a Bayesian colocalization test on the FUMA-annotated pleiotropic loci to identify potential shared causal variants in each pleiotropic locus. Colocalization analysis relies on single causal variant assumption and the posterior probability (PP) for five distinct hypotheses: (i) H_0_, no association within the under study region, (ii) H_1_, association only to the first trait, (iii) ­­H_2_, association only to the second trait, (iv) H_3_, association to both traits with distinct causal variants and (v) H_4_, associations to both traits with a shared causal variant[12]. Colocalization analyses were performed with the coloc.abf function from the coloc v.5.2.3 R package under the default settings. Genomic loci were declared as SNPs in ±200kb radius of the lead SNP as identified from FUMA. Loci with posterior probability (PP)H4>0.7 were identified as colocalized with a potential shared causal variant, that is the variant with the largest PP.H4 in the respective locus. Similar criteria were applied when examining all non-overlapping loci between AD and neuropsychiatric disorders, independently of statistical significance in each pairwise comparison. However, in the absence of a consensus lead variants at each independent locus, Bayesian colocalization was performed at the predefined loci.

The Multi-marker Analysis of GenoMic Annotation software (MAGMA) v1.10 was next used to perform gene and gene-set analyses[13]. The gene analysis was performed for SNPs around a ±10kb window around genes using the European reference panel of 1000 Genomes Project phase 3. We adopted the SNP-wide mean model for gene analyses. The summation of sample size in each pairwise trait was used as input for MAGMA v1.10. In the competitive gene-set analysis, we tested 17006 different gene-sets derived from MsigDB v2023.1Hs[14]. The results from both gene and gene-set analyses were corrected for multiple tests with the Bonferroni correction in a pairwise manner.

**Pleiotropy informing prioritization and evaluation**

To assess whether pleiotropic variants can be informative for drug repurposing opportunities, we used a recently proposed pipeline that (i) addresses the application of pleiotropy in drug discovery, (ii) provides a framework to evaluate the utility of pleiotropic evidence in target identification, and (iii) the extent to which pleiotropy can inform therapeutic strategies[15]. At first, pleiotropy informing prioritization and evaluation (PIPE) uses each pleiotropic variant to identify additional SNPs in strong LD (r^2^>0.8) using the European reference panel of the 1000 Genomes project phase 3. Each identified SNP is scored by using both LD strength and P-value associations. We submitted independent pleiotropic variants from each pairwise comparison in the PIPE pipeline, using all 24 Hi-C data and 69 eQTL datasets.

PIPE next leverages pleiotropic associations in a three-step manner, combining both molecular and high-quality network evidence. First, genes are prioritized according to genomic proximity (nGenes), chromatin interactions (cGenes) using 24 promoter Hi-C data and gene expression levels (eGenes) leveraging 69 eQTL datasets. An analytical list can be found on the corresponding publications[16-18]. Each gene is then scored according to the association of each SNP-gene pair for Hi-C and eQTL datasets in a cell- and tissue-specific manner, while nGene score depends on the collection of lead and LD SNPs within 20kb of each gene. Second, peripheral genes, that is genes connected to pleiotropic genes, are identified based on a random walk with restart algorithm using network evidence from experimental and database-related sources in the Search Tool for the Retrieval of Interacting Genes/Proteins (STRING) database[19], at a score cutoff of 0.7 from experimentally validated and database-documented interactions. A gene-predictor matrix is next formed with affinity scores according to (i) the relative importance of each predictor, that is genes whose removal results in significantly reduced model accuracy, and (ii) the importance of cGene and eGene predictors compared to the baseline nGene predictor representing the ground truth. In general, pleiotropic genes with increased connectivity degree to the ‘seed’ pleiotropic genes, that is the number of links in each node, receive higher affinity scores. The above affinity scores are finally converted to P-value-like scores and subsequently combined using Fisher’s combined probability test. The Fisher’s combined P-value is rescaled to 0-10 and represent the credit scores for each gene. Next, PIPE constructs a list of prioritized targets that likely mediate the pathway crosstalk between each pairwise comparison thus reflecting potential molecular mechanisms and highlighting relevant genes. Gene networks are constructed through the Kyoto Encyclopedia of Genes and Genomes (KEGG) database, as described previously, and are further denoted as ‘pathway crosstalks’.

The performance of PIPE in each trait pair is evaluated in a three-step manner. In specific, PIPE (i) computes the ability of informative predictors to distinguish between clinical proof-of-concept targets from simulated negative targets, (ii) quantifies the tendency of prioritized genes to be clinical proof-of-concept targets, and (iii) determining target-level disease relationships. Data regarding therapeutic approaches, clinical drug phases, target genes and relevant information were extracted from the ChEMBL database[23]. Simulated negative targets are constructed by excluding central and interacting[19] genes in each specific disease encompassing all druggable genes at any drug development phase. The performance of PIPE is measured through the area under the curve (AUC) metric, assessing the discriminative ability of each model to distinguish between clinical proof-of-concept targets in a disease-specific manner and simulated negative targets. Second, the leading prioritization analysis (LPA) aims to explore whether a gene list can be used as a clinical proof-of-concept target. The LPA follows a similar principle to gene set enrichment analysis using the prioritized gene list ranked by credit scores, where the enrichment score reflects whether a gene set is overrepresented at the extremes of a gene list[24]. Consequently, the PIPE algorithm computes the pleiotropy informing clinical therapeutics (PICT) metric, referring to the tendency of a prioritized gene list to contain clinical proof-of-concept targets, and depends on the fraction of clinical proof-of-concept targets recovered at the leading prioritization of the prioritized gene list. Third, inter-disease gene relationships are visualized as a network, with each gene, defined as a ‘member gene’ derived from the LPA analysis as a clinical proof-of-concept therapeutic target.

We next merged all five pathway crosstalks and performed a spin-glass model and simulated annealing clustering to identify discrete functional modules associated with AD and all neuropsychiatric disorders. In each module, we performed two discrete enrichment analyses to (i) describe biological pathways each module refers to using KEGG pathways, and (ii) disease enrichment analysis with DisGeneNet platform to identify modules relevant to AD. Overrepresentation analysis was performed to evaluate whether modules associated with AD were enriched for approved and phased drug targets.

Finally, we also examined the potential druggability of prioritized genes through constructing a supra-hexagonal map with the supraHex package[25]. For supraHex, we used as input the merged pathway crosstalks gene list containing credit scores for each pairwise comparison. Genes were classified as targetable if a known protein structure, derived from the protein databank (PDB) database[26], was predicted to contain druggable pockets through the fpocket software[27]. We divided the supra-hexagonal map in each identified cluster and color-coded according to credit scores in a pairwise comparison-specific manner. The cluster with the highest credit score among all pairwise comparisons was hypothesized to be strongly associated with AD contrary to comparison-specific clusters and was prioritized for further analysis. Genes in the cluster were visualized through the STRING v12.0 database using direct interactions at a high confidence score >0.7.

**Mendelian randomization**

We performed bi-directional Mendelian Randomization (MR) analyses to investigate the causal effects of genetic liability to neuropsychiatric disorders and AD and vice-versa. Variants selected as instrumental variables (IVs) should be (i) strongly associated with the exposure, (ii) independent of confounding factors in exposure-outcome association, and (iii) do not directly affect the outcome through other pathways. We selected independent SNPs reaching genome-wide significance (P-value<5×10^-8^) in each exposure as IVs. Instruments were clumped based on a local European 1000 Genomes Project phase 3 reference data using PLINK v1.9 within a 10Mb window, at an r^2^ threshold below 0.001. Outcome IVs were harmonized with the exposure variants to ensure that the effect estimate of a given SNP is oriented to the same allele. The mean F-statistic in each MR study was calculated, with F-statistic > 10 indicating an adequate instrument strength. In each outcome, we excluded variants with P-value<10^-4^ to reduce pleiotropic effects.

The inverse-variance weighted (IVW) method served as our primary outcome. The Cochran’s Q statistic was used to estimate the level of heterogeneity, quantified with the Higgins *I^2^* statistic[28]. Complementary sensitivity analyses were conducted to assess the robustness of our MR results, such as the leave-one-SNP-out analysis, MR-Egger and weighted median. Weighted median provides consistent estimates when more than 50% of the total weight is contributed by the selected IVs[29]. MR-Egger is an adaptation of IVW that captures the weighted average pleiotropic effect by introducing an intercept term[30]. The MR pleiotropy residual sum and outlier (MR-PRESSO) method was used to investigate potential outliers, by (i) detecting the presence of horizontal pleiotropy, (ii) correcting the causal estimate via removing outliers and (iii) conducting a distortion test to evaluate differences in the causal estimates pre- and post-outlier correction[31]. In cases of sample overlap as reported from the corresponding GWAS publications, we applied MRlap[32], a recently proposed method that accounts for sample overlap using GWAS summary statistics by employing cross-trait LDSC[7]. The MRlap pipeline requires sample overlap independent of case-control status, as well as the total sample size (ncases + ncontrols). All reported estimates from the MRlap are reported per SD scale. For MRlap, we excluded IVs identified as outliers from MR-PRESSO. If the estimates of MRlap differ substantially compared to the observed effect, as quantified with test statistic, then the MR-IVW corrected effects should be preferred.

**Supplementary references**

1. Budu-Aggrey, A. Kilanowski, M. K. Sobczyk, 23andMe Research Team, S. S. Shringarpure, R. Mitchell, K. Reis, A. Reigo, Estonian Biobank Research Team, R. Mägi, M. Nelis, N. Tanaka, B. M. Brumpton, L. F. Thomas, P. Sole-Navais, C. Flatley, A. Espuela-Ortiz, E. Herrera-Luis, J. V. T. Lominchar, J. Bork-Jensen, I. Marenholz, A. Arnau-Soler, A. Jeong, K. A. Fawcett, H. Baurecht, E. Rodriguez, A. C. Alves, A. Kumar, P. M. Sleiman, X. Chang, C. Medina-Gomez, C. Hu, C. Xu, C. Qi, S. El-Heis, P. Titcombe, E. Antoun, J. Fadista, C. A. Wang, E. Thiering, B. Wu, S. Kress, D. M. Kothalawala, L. Kadalayil, J. Duan, H. Zhang, S. Hadebe, T. Hoffmann, E. Jorgenson, H. Choquet, N. Risch, P. Njølstad, O. A. Andreassen, S. Johansson, C. Almqvist, T. Gong, V. Ullemar, R. Karlsson, P. K. E. Magnusson, A. Szwajda, E. G. Burchard, J. P. Thyssen, T. Hansen, L. L. Kårhus, T. M. Dantoft, A. C. S. N. Jeanrenaud, A. Ghauri, A. Arnold, G. Homuth, S. Lau, M. M. Nöthen, N. Hübner, M. Imboden, A. Visconti, M. Falchi, V. Bataille, P. Hysi, N. Ballardini, D. I. Boomsma, J. J. Hottenga, M. Müller-Nurasyid, T. S. Ahluwalia, J. Stokholm, B. Chawes, A.-M. M. Schoos, A. Esplugues, M. Bustamante, B. Raby, S. Arshad, C. German, T. Esko, L. A. Milani, A. Metspalu, C. Terao, K. Abuabara, M. Løset, K. Hveem, B. Jacobsson, M. Pino-Yanes, D. P. Strachan, N. Grarup, A. Linneberg, Y.-A. Lee, N. Probst-Hensch, S. Weidinger, M.-R. Jarvelin, E. Melén, H. Hakonarson, A. D. Irvine, D. Jarvis, T. Nijsten, L. Duijts, J. M. Vonk, G. H. Koppelmann, K. M. Godfrey, S. J. Barton, B. Feenstra, C. E. Pennell, P. D. Sly, P. G. Holt, L. K. Williams, H. Bisgaard, K. Bønnelykke, J. Curtin, A. Simpson, C. Murray, T. Schikowski, S. Bunyavanich, S. T. Weiss, J. W. Holloway, J. L. Min, S. J. Brown, M. Standl, L. Paternoster, European and multi-ancestry genome-wide association meta-analysis of atopic dermatitis highlights importance of systemic immune regulation. *Nat Commun* **14**, 6172 (2023).
2. D. Demontis, G. B. Walters, G. Athanasiadis, R. Walters, K. Therrien, T. T. Nielsen, L. Farajzadeh, G. Voloudakis, J. Bendl, B. Zeng, W. Zhang, J. Grove, T. D. Als, J. Duan, F. K. Satterstrom, J. Bybjerg-Grauholm, M. Bækved-Hansen, O. O. Gudmundsson, S. H. Magnusson, G. Baldursson, K. Davidsdottir, G. S. Haraldsdottir, E. Agerbo, G. E. Hoffman, S. Dalsgaard, J. Martin, M. Ribasés, D. I. Boomsma, M. Soler Artigas, N. Roth Mota, D. Howrigan, S. E. Medland, T. Zayats, V. M. Rajagopal, ADHD Working Group of the Psychiatric Genomics Consortium, A. Havdahl, A. Doyle, A. Reif, A. Thapar, B. Cormand, C. Liao, C. Burton, C. H. D. Bau, D. L. Rovaris, E. Sonuga-Barke, E. Corfield, E. H. Grevet, H. Larsson, I. R. Gizer, I. Waldman, I. Brikell, J. Haavik, J. Crosbie, J. McGough, J. Kuntsi, J. Glessner, K. Langley, K.-P. Lesch, L. A. Rohde, M. H. Hutz, M. Klein, M. Bellgrove, M. Tesli, M. C. O’Donovan, O. A. Andreassen, P. W. L. Leung, P. M. Pan, R. Joober, R. Schachar, S. Loo, S. H. Witt, T. Reichborn-Kjennerud, T. Banaschewski, Z. Hawi, iPSYCH-Broad Consortium, M. J. Daly, O. Mors, M. Nordentoft, O. Mors, D. M. Hougaard, P. B. Mortensen, M. J. Daly, S. V. Faraone, H. Stefansson, P. Roussos, B. Franke, T. Werge, B. M. Neale, K. Stefansson, A. D. Børglum, Genome-wide analyses of ADHD identify 27 risk loci, refine the genetic architecture and implicate several cognitive domains. *Nat Genet* **55**, 198–208 (2023).
3. Autism Spectrum Disorder Working Group of the Psychiatric Genomics Consortium, BUPGEN, Major Depressive Disorder Working Group of the Psychiatric Genomics Consortium, 23andMe Research Team, J. Grove, S. Ripke, T. D. Als, M. Mattheisen, R. K. Walters, H. Won, J. Pallesen, E. Agerbo, O. A. Andreassen, R. Anney, S. Awashti, R. Belliveau, F. Bettella, J. D. Buxbaum, J. Bybjerg-Grauholm, M. Bækvad-Hansen, F. Cerrato, K. Chambert, J. H. Christensen, C. Churchhouse, K. Dellenvall, D. Demontis, S. De Rubeis, B. Devlin, S. Djurovic, A. L. Dumont, J. I. Goldstein, C. S. Hansen, M. E. Hauberg, M. V. Hollegaard, S. Hope, D. P. Howrigan, H. Huang, C. M. Hultman, L. Klei, J. Maller, J. Martin, A. R. Martin, J. L. Moran, M. Nyegaard, T. Nærland, D. S. Palmer, A. Palotie, C. B. Pedersen, M. G. Pedersen, T. dPoterba, J. B. Poulsen, B. S. Pourcain, P. Qvist, K. Rehnström, A. Reichenberg, J. Reichert, E. B. Robinson, K. Roeder, P. Roussos, E. Saemundsen, S. Sandin, F. K. Satterstrom, G. Davey Smith, H. Stefansson, S. Steinberg, C. R. Stevens, P. F. Sullivan, P. Turley, G. B. Walters, X. Xu, K. Stefansson, D. H. Geschwind, M. Nordentoft, D. M. Hougaard, T. Werge, O. Mors, P. B. Mortensen, B. M. Neale, M. J. Daly, A. D. Børglum, Identification of common genetic risk variants for autism spectrum disorder. *Nat Genet* **51**, 431–444 (2019).
4. N. Mullins, A. J. Forstner, K. S. O’Connell, B. Coombes, J. R. I. Coleman, Z. Qiao, T. D. Als, T. B. Bigdeli, S. Børte, J. Bryois, A. W. Charney, O. K. Drange, M. J. Gandal, S. P. Hagenaars, M. Ikeda, N. Kamitaki, M. Kim, K. Krebs, G. Panagiotaropoulou, B. M. Schilder, L. G. Sloofman, S. Steinberg, V. Trubetskoy, B. S. Winsvold, H.-H. Won, L. Abramova, K. Adorjan, E. Agerbo, M. Al Eissa, D. Albani, N. Alliey-Rodriguez, A. Anjorin, V. Antilla, A. Antoniou, S. Awasthi, J. H. Baek, M. Bækvad-Hansen, N. Bass, M. Bauer, E. C. Beins, S. E. Bergen, A. Birner, C. Bøcker Pedersen, E. Bøen, M. P. Boks, R. Bosch, M. Brum, B. M. Brumpton, N. Brunkhorst-Kanaan, M. Budde, J. Bybjerg-Grauholm, W. Byerley, M. Cairns, M. Casas, P. Cervantes, T.-K. Clarke, C. Cruceanu, A. Cuellar-Barboza, J. Cunningham, D. Curtis, P. M. Czerski, A. M. Dale, N. Dalkner, F. S. David, F. Degenhardt, S. Djurovic, A. L. Dobbyn, A. Douzenis, T. Elvsåshagen, V. Escott-Price, I. N. Ferrier, A. Fiorentino, T. M. Foroud, L. Forty, J. Frank, O. Frei, N. B. Freimer, L. Frisén, K. Gade, J. Garnham, J. Gelernter, M. Giørtz Pedersen, I. R. Gizer, S. D. Gordon, K. Gordon-Smith, T. A. Greenwood, J. Grove, J. Guzman-Parra, K. Ha, M. Haraldsson, M. Hautzinger, U. Heilbronner, D. Hellgren, S. Herms, P. Hoffmann, P. A. Holmans, L. Huckins, S. Jamain, J. S. Johnson, J. L. Kalman, Y. Kamatani, J. L. Kennedy, S. Kittel-Schneider, J. A. Knowles, M. Kogevinas, M. Koromina, T. M. Kranz, H. R. Kranzler, M. Kubo, R. Kupka, S. A. Kushner, C. Lavebratt, J. Lawrence, M. Leber, H.-J. Lee, P. H. Lee, S. E. Levy, C. Lewis, C. Liao, S. Lucae, M. Lundberg, D. J. MacIntyre, S. H. Magnusson, W. Maier, A. Maihofer, D. Malaspina, E. Maratou, L. Martinsson, M. Mattheisen, S. A. McCarroll, N. W. McGregor, P. McGuffin, J. D. McKay, H. Medeiros, S. E. Medland, V. Millischer, G. W. Montgomery, J. L. Moran, D. W. Morris, T. W. Mühleisen, N. O’Brien, C. O’Donovan, L. M. Olde Loohuis, L. Oruc, S. Papiol, A. F. Pardiñas, A. Perry, A. Pfennig, E. Porichi, J. B. Potash, D. Quested, T. Raj, M. H. Rapaport, J. R. DePaulo, E. J. Regeer, J. P. Rice, F. Rivas, M. Rivera, J. Roth, P. Roussos, D. M. Ruderfer, C. Sánchez-Mora, E. C. Schulte, F. Senner, S. Sharp, P. D. Shilling, E. Sigurdsson, L. Sirignano, C. Slaney, O. B. Smeland, D. J. Smith, J. L. Sobell, C. Søholm Hansen, M. Soler Artigas, A. T. Spijker, D. J. Stein, J. S. Strauss, B. Świątkowska, C. Terao, T. E. Thorgeirsson, C. Toma, P. Tooney, E.-E. Tsermpini, M. P. Vawter, H. Vedder, J. T. R. Walters, S. H. Witt, S. Xi, W. Xu, J. M. K. Yang, A. H. Young, H. Young, P. P. Zandi, H. Zhou, L. Zillich, HUNT All-In Psychiatry, R. Adolfsson, I. Agartz, M. Alda, L. Alfredsson, G. Babadjanova, L. Backlund, B. T. Baune, F. Bellivier, S. Bengesser, W. H. Berrettini, D. H. R. Blackwood, M. Boehnke, A. D. Børglum, G. Breen, V. J. Carr, S. Catts, A. Corvin, N. Craddock, U. Dannlowski, D. Dikeos, T. Esko, B. Etain, P. Ferentinos, M. Frye, J. M. Fullerton, M. Gawlik, E. S. Gershon, F. S. Goes, M. J. Green, M. Grigoroiu-Serbanescu, J. Hauser, F. Henskens, J. Hillert, K. S. Hong, D. M. Hougaard, C. M. Hultman, K. Hveem, N. Iwata, A. V. Jablensky, I. Jones, L. A. Jones, R. S. Kahn, J. R. Kelsoe, G. Kirov, M. Landén, M. Leboyer, C. M. Lewis, Q. S. Li, J. Lissowska, C. Lochner, C. Loughland, N. G. Martin, C. A. Mathews, F. Mayoral, S. L. McElroy, A. M. McIntosh, F. J. McMahon, I. Melle, P. Michie, L. Milani, P. B. Mitchell, G. Morken, O. Mors, P. B. Mortensen, B. Mowry, B. Müller-Myhsok, R. M. Myers, B. M. Neale, C. M. Nievergelt, M. Nordentoft, M. M. Nöthen, M. C. O’Donovan, K. J. Oedegaard, T. Olsson, M. J. Owen, S. A. Paciga, C. Pantelis, C. Pato, M. T. Pato, G. P. Patrinos, R. H. Perlis, D. Posthuma, J. A. Ramos-Quiroga, A. Reif, E. Z. Reininghaus, M. Ribasés, M. Rietschel, S. Ripke, G. A. Rouleau, T. Saito, U. Schall, M. Schalling, P. R. Schofield, T. G. Schulze, L. J. Scott, R. J. Scott, A. Serretti, C. Shannon Weickert, J. W. Smoller, H. Stefansson, K. Stefansson, E. Stordal, F. Streit, P. F. Sullivan, G. Turecki, A. E. Vaaler, E. Vieta, J. B. Vincent, I. D. Waldman, T. W. Weickert, T. Werge, N. R. Wray, J.-A. Zwart, J. M. Biernacka, J. I. Nurnberger, S. Cichon, H. J. Edenberg, E. A. Stahl, A. McQuillin, A. Di Florio, R. A. Ophoff, O. A. Andreassen, Genome-wide association study of more than 40,000 bipolar disorder cases provides new insights into the underlying biology. *Nat Genet* **53**, 817–829 (2021).
5. D. M. Howard, M. J. Adams, T.-K. Clarke, J. D. Hafferty, J. Gibson, M. Shirali, J. R. I. Coleman, S. P. Hagenaars, J. Ward, E. M. Wigmore, C. Alloza, X. Shen, M. C. Barbu, E. Y. Xu, H. C. Whalley, R. E. Marioni, D. J. Porteous, G. Davies, I. J. Deary, G. Hemani, K. Berger, H. Teismann, R. Rawal, V. Arolt, B. T. Baune, U. Dannlowski, K. Domschke, C. Tian, D. A. Hinds, 23andMe Research Team, Major Depressive Disorder Working Group of the Psychiatric Genomics Consortium, M. Trzaskowski, E. M. Byrne, S. Ripke, D. J. Smith, P. F. Sullivan, N. R. Wray, G. Breen, C. M. Lewis, A. M. McIntosh, Genome-wide meta-analysis of depression identifies 102 independent variants and highlights the importance of the prefrontal brain regions. *Nat Neurosci* **22**, 343–352 (2019).
6. V. Trubetskoy, A. F. Pardiñas, T. Qi, G. Panagiotaropoulou, S. Awasthi, T. B. Bigdeli, J. Bryois, C.-Y. Chen, C. A. Dennison, L. S. Hall, M. Lam, K. Watanabe, O. Frei, T. Ge, J. C. Harwood, F. Koopmans, S. Magnusson, A. L. Richards, J. Sidorenko, Y. Wu, J. Zeng, J. Grove, M. Kim, Z. Li, G. Voloudakis, W. Zhang, M. Adams, I. Agartz, E. G. Atkinson, E. Agerbo, M. Al Eissa, M. Albus, M. Alexander, B. Z. Alizadeh, K. Alptekin, T. D. Als, F. Amin, V. Arolt, M. Arrojo, L. Athanasiu, M. H. Azevedo, S. A. Bacanu, N. J. Bass, M. Begemann, R. A. Belliveau, J. Bene, B. Benyamin, S. E. Bergen, G. Blasi, J. Bobes, S. Bonassi, A. Braun, R. A. Bressan, E. J. Bromet, R. Bruggeman, P. F. Buckley, R. L. Buckner, J. Bybjerg-Grauholm, W. Cahn, M. J. Cairns, M. E. Calkins, V. J. Carr, D. Castle, S. V. Catts, K. D. Chambert, R. C. K. Chan, B. Chaumette, W. Cheng, E. F. C. Cheung, S. A. Chong, D. Cohen, A. Consoli, Q. Cordeiro, J. Costas, C. Curtis, M. Davidson, K. L. Davis, L. De Haan, F. Degenhardt, L. E. DeLisi, D. Demontis, F. Dickerson, D. Dikeos, T. Dinan, S. Djurovic, J. Duan, G. Ducci, F. Dudbridge, J. G. Eriksson, L. Fañanás, S. V. Faraone, A. Fiorentino, A. Forstner, J. Frank, N. B. Freimer, M. Fromer, A. Frustaci, A. Gadelha, G. Genovese, E. S. Gershon, M. Giannitelli, I. Giegling, P. Giusti-Rodríguez, S. Godard, J. I. Goldstein, J. González Peñas, A. González-Pinto, S. Gopal, J. Gratten, M. F. Green, T. A. Greenwood, O. Guillin, S. Gülöksüz, R. E. Gur, R. C. Gur, B. Gutiérrez, E. Hahn, H. Hakonarson, V. Haroutunian, A. M. Hartmann, C. Harvey, C. Hayward, F. A. Henskens, S. Herms, P. Hoffmann, D. P. Howrigan, M. Ikeda, C. Iyegbe, I. Joa, A. Julià, A. K. Kähler, T. Kam-Thong, Y. Kamatani, S. Karachanak-Yankova, O. Kebir, M. C. Keller, B. J. Kelly, A. Khrunin, S.-W. Kim, J. Klovins, N. Kondratiev, B. Konte, J. Kraft, M. Kubo, V. Kučinskas, Z. A. Kučinskiene, A. Kusumawardhani, H. Kuzelova-Ptackova, S. Landi, L. C. Lazzeroni, P. H. Lee, S. E. Legge, D. S. Lehrer, R. Lencer, B. Lerer, M. Li, J. Lieberman, G. A. Light, S. Limborska, C.-M. Liu, J. Lönnqvist, C. M. Loughland, J. Lubinski, J. J. Luykx, A. Lynham, M. Macek, A. Mackinnon, P. K. E. Magnusson, B. S. Maher, W. Maier, D. Malaspina, J. Mallet, S. R. Marder, S. Marsal, A. R. Martin, L. Martorell, M. Mattheisen, R. W. McCarley, C. McDonald, J. J. McGrath, H. Medeiros, S. Meier, B. Melegh, I. Melle, R. I. Mesholam-Gately, A. Metspalu, P. T. Michie, L. Milani, V. Milanova, M. Mitjans, E. Molden, E. Molina, M. D. Molto, V. Mondelli, C. Moreno, C. P. Morley, G. Muntané, K. C. Murphy, I. Myin-Germeys, I. Nenadić, G. Nestadt, L. Nikitina-Zake, C. Noto, K. H. Nuechterlein, N. L. O’Brien, F. A. O’Neill, S.-Y. Oh, A. Olincy, V. K. Ota, C. Pantelis, G. N. Papadimitriou, M. Parellada, T. Paunio, R. Pellegrino, S. Periyasamy, D. O. Perkins, B. Pfuhlmann, O. Pietiläinen, J. Pimm, D. Porteous, J. Powell, D. Quattrone, D. Quested, A. D. Radant, A. Rampino, M. H. Rapaport, A. Rautanen, A. Reichenberg, C. Roe, J. L. Roffman, J. Roth, M. Rothermundt, B. P. F. Rutten, S. Saker-Delye, V. Salomaa, J. Sanjuan, M. L. Santoro, A. Savitz, U. Schall, R. J. Scott, L. J. Seidman, S. I. Sharp, J. Shi, L. J. Siever, E. Sigurdsson, K. Sim, N. Skarabis, P. Slominsky, H.-C. So, J. L. Sobell, E. Söderman, H. J. Stain, N. E. Steen, A. A. Steixner-Kumar, E. Stögmann, W. S. Stone, R. E. Straub, F. Streit, E. Strengman, T. S. Stroup, M. Subramaniam, C. A. Sugar, J. Suvisaari, D. M. Svrakic, N. R. Swerdlow, J. P. Szatkiewicz, T. M. T. Ta, A. Takahashi, C. Terao, F. Thibaut, D. Toncheva, P. A. Tooney, S. Torretta, S. Tosato, G. B. Tura, B. I. Turetsky, A. Üçok, A. Vaaler, T. Van Amelsvoort, R. Van Winkel, J. Veijola, J. Waddington, H. Walter, A. Waterreus, B. T. Webb, M. Weiser, N. M. Williams, S. H. Witt, B. K. Wormley, J. Q. Wu, Z. Xu, R. Yolken, C. C. Zai, W. Zhou, F. Zhu, F. Zimprich, E. C. Atbaşoğlu, M. Ayub, C. Benner, A. Bertolino, D. W. Black, N. J. Bray, G. Breen, N. G. Buccola, W. F. Byerley, W. J. Chen, C. R. Cloninger, B. Crespo-Facorro, G. Donohoe, R. Freedman, C. Galletly, M. J. Gandal, M. Gennarelli, D. M. Hougaard, H.-G. Hwu, A. V. Jablensky, S. A. McCarroll, J. L. Moran, O. Mors, P. B. Mortensen, B. Müller-Myhsok, A. L. Neil, M. Nordentoft, M. T. Pato, T. L. Petryshen, M. Pirinen, A. E. Pulver, T. G. Schulze, J. M. Silverman, J. W. Smoller, E. A. Stahl, D. W. Tsuang, E. Vilella, S.-H. Wang, S. Xu, Indonesia Schizophrenia Consortium, N. Dai, Q. Wenwen, D. B. Wildenauer, F. Agiananda, N. Amir, R. Antoni, T. Arsianti, A. Asmarahadi, H. Diatri, P. Djatmiko, I. Irmansyah, S. Khalimah, I. Kusumadewi, P. Kusumaningrum, P. R. Lukman, M. W. Nasrun, N. S. Safyuni, P. Prasetyawan, G. Semen, K. Siste, H. Tobing, N. Widiasih, T. Wiguna, D. Wulandari, N. Evalina, A. J. Hananto, J. H. Ismoyo, T. M. Marini, S. Henuhili, M. Reza, S. Yusnadewi, PsychENCODE, A. Abyzov, S. Akbarian, A. Ashley-Koch, H. Van Bakel, M. Breen, M. Brown, J. Bryois, B. Carlyle, A. Charney, G. Coetzee, G. Crawford, S. Dracheva, P. Emani, P. Farnham, M. Fromer, T. Galeev, M. Gandal, M. Gerstein, G. Giase, K. Girdhar, F. Goes, K. Grennan, M. Gu, B. Guerra, G. Gursoy, G. Hoffman, T. Hyde, A. Jaffe, S. Jiang, Y. Jiang, A. Kefi, Y. Kim, R. Kitchen, J. A. Knowles, F. Lay, D. Lee, M. Li, C. Liu, S. Liu, E. Mattei, F. Navarro, X. Pan, M. A. Peters, D. Pinto, S. Pochareddy, D. Polioudakis, M. Purcaro, S. Purcell, H. Pratt, T. Reddy, S. Rhie, P. Roussos, J. Rozowsky, S. Sanders, N. Sestan, A. Sethi, X. Shi, A. Shieh, V. Swarup, A. Szekely, D. Wang, J. Warrell, S. Weissman, Z. Weng, K. White, J. Wiseman, H. Witt, H. Won, S. Wood, F. Wu, X. Xu, L. Yao, P. Zandi, Psychosis Endophenotypes International Consortium, M. J. Arranz, S. Bakker, S. Bender, E. Bramon, D. A. Collier, B. Crepo-Facorro, J. Hall, C. Iyegbe, R. Kahn, S. Lawrie, C. Lewis, K. Lin, D. H. Linszen, I. Mata, A. McIntosh, R. M. Murray, R. A. Ophoff, J. Van Os, J. Powell, D. Rujescu, M. Walshe, M. Weisbrod, The SynGO Consortium, T. Achsel, M. Andres-Alonso, C. Bagni, À. Bayés, T. Biederer, N. Brose, T. C. Brown, J. J. E. Chua, M. P. Coba, L. N. Cornelisse, A. P. H. De Jong, J. De Juan-Sanz, D. C. Dieterich, G. Feng, H. L. Goldschmidt, E. D. Gundelfinger, C. Hoogenraad, R. L. Huganir, S. E. Hyman, C. Imig, R. Jahn, H. Jung, P. S. Kaeser, E. Kim, F. Koopmans, M. R. Kreutz, N. Lipstein, H. D. MacGillavry, R. Malenka, P. S. McPherson, V. O’Connor, R. Pielot, T. A. Ryan, D. Sahasrabudhe, C. Sala, M. Sheng, K.-H. Smalla, A. B. Smit, T. C. Südhof, P. D. Thomas, R. F. Toonen, J. R. T. Van Weering, M. Verhage, C. Verpelli, R. Adolfsson, C. Arango, B. T. Baune, S. I. Belangero, A. D. Børglum, D. Braff, E. Bramon, J. D. Buxbaum, D. Campion, J. A. Cervilla, S. Cichon, D. A. Collier, A. Corvin, D. Curtis, M. D. Forti, E. Domenici, H. Ehrenreich, V. Escott-Price, T. Esko, A. H. Fanous, A. Gareeva, M. Gawlik, P. V. Gejman, M. Gill, S. J. Glatt, V. Golimbet, K. S. Hong, C. M. Hultman, S. E. Hyman, N. Iwata, E. G. Jönsson, R. S. Kahn, J. L. Kennedy, E. Khusnutdinova, G. Kirov, J. A. Knowles, M.-O. Krebs, C. Laurent-Levinson, J. Lee, T. Lencz, D. F. Levinson, Q. S. Li, J. Liu, A. K. Malhotra, D. Malhotra, A. McIntosh, A. McQuillin, P. R. Menezes, V. A. Morgan, D. W. Morris, B. J. Mowry, R. M. Murray, V. Nimgaonkar, M. M. Nöthen, R. A. Ophoff, S. A. Paciga, A. Palotie, C. N. Pato, S. Qin, M. Rietschel, B. P. Riley, M. Rivera, D. Rujescu, M. C. Saka, A. R. Sanders, S. G. Schwab, A. Serretti, P. C. Sham, Y. Shi, D. St Clair, H. Stefánsson, K. Stefansson, M. T. Tsuang, J. Van Os, M. P. Vawter, D. R. Weinberger, T. Werge, D. B. Wildenauer, X. Yu, W. Yue, P. A. Holmans, A. J. Pocklington, P. Roussos, E. Vassos, M. Verhage, P. M. Visscher, J. Yang, D. Posthuma, O. A. Andreassen, K. S. Kendler, M. J. Owen, N. R. Wray, M. J. Daly, H. Huang, B. M. Neale, P. F. Sullivan, S. Ripke, J. T. R. Walters, M. C. O’Donovan, Schizophrenia Working Group of the Psychiatric Genomics Consortium, L. De Haan, T. Van Amelsvoort, R. Van Winkel, A. Gareeva, P. C. Sham, Y. Shi, D. St Clair, J. Van Os, Mapping genomic loci implicates genes and synaptic biology in schizophrenia. *Nature* **604**, 502–508 (2022).
7. Schizophrenia Working Group of the Psychiatric Genomics Consortium, B. K. Bulik-Sullivan, P.-R. Loh, H. K. Finucane, S. Ripke, J. Yang, N. Patterson, M. J. Daly, A. L. Price, B. M. Neale, LD Score regression distinguishes confounding from polygenicity in genome-wide association studies. *Nat Genet* **47**, 291–295 (2015).
8. D. Ray, N. Chatterjee, A powerful method for pleiotropic analysis under composite null hypothesis identifies novel shared loci between Type 2 Diabetes and Prostate Cancer. *PLoS Genet* **16**, e1009218 (2020).
9. K. Wang, M. Li, H. Hakonarson, ANNOVAR: functional annotation of genetic variants from high-throughput sequencing data. *Nucleic Acids Research* **38**, e164–e164 (2010).
10. K. Watanabe, E. Taskesen, A. Van Bochoven, D. Posthuma, Functional mapping and annotation of genetic associations with FUMA. *Nat Commun* **8**, 1826 (2017).
11. International Inflammatory Bowel Disease Genetics Consortium, H. Huang, M. Fang, L. Jostins, M. Umićević Mirkov, G. Boucher, C. A. Anderson, V. Andersen, I. Cleynen, A. Cortes, F. Crins, M. D’Amato, V. Deffontaine, J. Dmitrieva, E. Docampo, M. Elansary, K. K.-H. Farh, A. Franke, A.-S. Gori, P. Goyette, J. Halfvarson, T. Haritunians, J. Knight, I. C. Lawrance, C. W. Lees, E. Louis, R. Mariman, T. Meuwissen, M. Mni, Y. Momozawa, M. Parkes, S. L. Spain, E. Théâtre, G. Trynka, J. Satsangi, S. Van Sommeren, S. Vermeire, R. J. Xavier, R. K. Weersma, R. H. Duerr, C. G. Mathew, J. D. Rioux, D. P. B. McGovern, J. H. Cho, M. Georges, M. J. Daly, J. C. Barrett, Fine-mapping inflammatory bowel disease loci to single-variant resolution. *Nature* **547**, 173–178 (2017).
12. C. Giambartolomei, D. Vukcevic, E. E. Schadt, L. Franke, A. D. Hingorani, C. Wallace, V. Plagnol, Bayesian Test for Colocalisation between Pairs of Genetic Association Studies Using Summary Statistics. *PLoS Genet* **10**, e1004383 (2014).
13. C. A. De Leeuw, J. M. Mooij, T. Heskes, D. Posthuma, MAGMA: Generalized Gene-Set Analysis of GWAS Data. *PLoS Comput Biol* **11**, e1004219 (2015).
14. A. Liberzon, C. Birger, H. Thorvaldsdóttir, M. Ghandi, J. P. Mesirov, P. Tamayo, The Molecular Signatures Database Hallmark Gene Set Collection. *Cell Systems* **1**, 417–425 (2015).
15. C. Bao, T. Tan, S. Wang, C. Gao, C. Lu, S. Yang, Y. Diao, L. Jiang, D. Jing, L. Chen, H. Lv, H. Fang, A cross-disease, pleiotropy-driven approach for therapeutic target prioritization and evaluation. *Cell Reports Methods* **4**, 100757 (2024).
16. I. Jung, A. Schmitt, Y. Diao, A. J. Lee, T. Liu, D. Yang, C. Tan, J. Eom, M. Chan, S. Chee, Z. Chiang, C. Kim, E. Masliah, C. L. Barr, B. Li, S. Kuan, D. Kim, B. Ren, A compendium of promoter-centered long-range chromatin interactions in the human genome. *Nat Genet* **51**, 1442–1449 (2019).
17. M. Song, X. Yang, X. Ren, L. Maliskova, B. Li, I. R. Jones, C. Wang, F. Jacob, K. Wu, M. Traglia, T. W. Tam, K. Jamieson, S.-Y. Lu, G.-L. Ming, Y. Li, J. Yao, L. A. Weiss, J. R. Dixon, L. M. Judge, B. R. Conklin, H. Song, L. Gan, Y. Shen, Mapping cis-regulatory chromatin contacts in neural cells links neuropsychiatric disorder risk variants to target genes. *Nat Genet* **51**, 1252–1262 (2019).
18. N. Kerimov, J. D. Hayhurst, K. Peikova, J. R. Manning, P. Walter, L. Kolberg, M. Samoviča, M. P. Sakthivel, I. Kuzmin, S. J. Trevanion, T. Burdett, S. Jupp, H. Parkinson, I. Papatheodorou, A. D. Yates, D. R. Zerbino, K. Alasoo, A compendium of uniformly processed human gene expression and splicing quantitative trait loci. *Nat Genet* **53**, 1290–1299 (2021).
19. D. Szklarczyk, R. Kirsch, M. Koutrouli, K. Nastou, F. Mehryary, R. Hachilif, A. L. Gable, T. Fang, N. T. Doncheva, S. Pyysalo, P. Bork, L. J. Jensen, C. von Mering, The STRING database in 2023: protein–protein association networks and functional enrichment analyses for any sequenced genome of interest. *Nucleic Acids Research* **51**, D638–D646 (2023).
20. H. Fang, ULTRA-DD Consortium, H. De Wolf, B. Knezevic, K. L. Burnham, J. Osgood, A. Sanniti, A. Lledó Lara, S. Kasela, S. De Cesco, J. K. Wegner, L. Handunnetthi, F. E. McCann, L. Chen, T. Sekine, P. E. Brennan, B. D. Marsden, D. Damerell, C. A. O’Callaghan, C. Bountra, P. Bowness, Y. Sundström, L. Milani, L. Berg, H. W. Göhlmann, P. J. Peeters, B. P. Fairfax, M. Sundström, J. C. Knight, A genetics-led approach defines the drug target landscape of 30 immune-related traits. *Nat Genet* **51**, 1082–1091 (2019).
21. M. Kanehisa, M. Furumichi, Y. Sato, M. Kawashima, M. Ishiguro-Watanabe, KEGG for taxonomy-based analysis of pathways and genomes. *Nucleic Acids Res* **51**, D587–D592 (2023).
22. H. Fang, J. Gough, The “dnet” approach promotes emerging research on cancer patient survival. *Genome Med* **6**, 64 (2014).
23. B. Zdrazil, E. Felix, F. Hunter, E. J. Manners, J. Blackshaw, S. Corbett, M. de Veij, H. Ioannidis, D. M. Lopez, J. F. Mosquera, M. P. Magarinos, N. Bosc, R. Arcila, T. Kizilören, A. Gaulton, A. P. Bento, M. F. Adasme, P. Monecke, G. A. Landrum, A. R. Leach, The ChEMBL Database in 2023: a drug discovery platform spanning multiple bioactivity data types and time periods. *Nucleic Acids Res* **52**, D1180–D1192 (2024).
24. A. Subramanian, P. Tamayo, V. K. Mootha, S. Mukherjee, B. L. Ebert, M. A. Gillette, A. Paulovich, S. L. Pomeroy, T. R. Golub, E. S. Lander, J. P. Mesirov, Gene set enrichment analysis: A knowledge-based approach for interpreting genome-wide expression profiles. *Proc. Natl. Acad. Sci. U.S.A.* **102**, 15545–15550 (2005).
25. H. Fang, J. Gough, supraHex: an R/Bioconductor package for tabular omics data analysis using a supra-hexagonal map. *Biochem Biophys Res Commun* **443**, 285–289 (2014).
26. H. M. Berman, The Protein Data Bank. *Nucleic Acids Research* **28**, 235–242 (2000).
27. V. Le Guilloux, P. Schmidtke, P. Tuffery, Fpocket: An open source platform for ligand pocket detection. *BMC Bioinformatics* **10**, 168 (2009).
28. J. P. T. Higgins, S. G. Thompson, J. J. Deeks, D. G. Altman, Measuring inconsistency in meta-analyses. *BMJ* **327**, 557–560 (2003).
29. J. Bowden, G. Davey Smith, P. C. Haycock, S. Burgess, Consistent Estimation in Mendelian Randomization with Some Invalid Instruments Using a Weighted Median Estimator. *Genet Epidemiol* **40**, 304–314 (2016).
30. J. Bowden, G. Davey Smith, S. Burgess, Mendelian randomization with invalid instruments: effect estimation and bias detection through Egger regression. *Int J Epidemiol* **44**, 512–525 (2015).
31. M. Verbanck, C.-Y. Chen, B. Neale, R. Do, Detection of widespread horizontal pleiotropy in causal relationships inferred from Mendelian randomization between complex traits and diseases. *Nat Genet* **50**, 693–698 (2018).
32. N. Mounier, Z. Kutalik, Bias correction for inverse variance weighting Mendelian randomization. *Genetic Epidemiology* **47**, 314–331 (2023).
