## Supplementary Figures for "A genome-wide pleiotropy study between atopic dermatitis and neuropsychiatric disorders"

**Charalabos Antonatos *et al*.**

Fig. S1. Quantile-Quantile (QQ) plots of all pairwise comparisons.

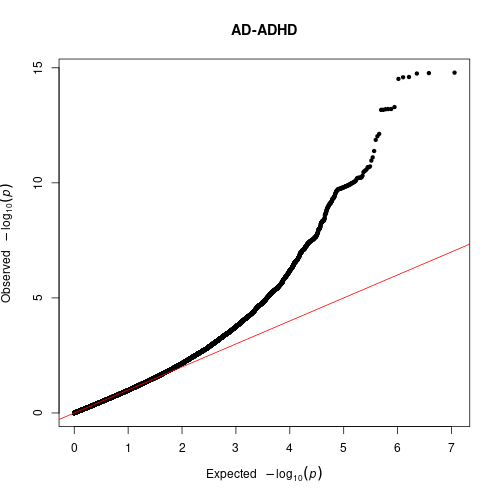

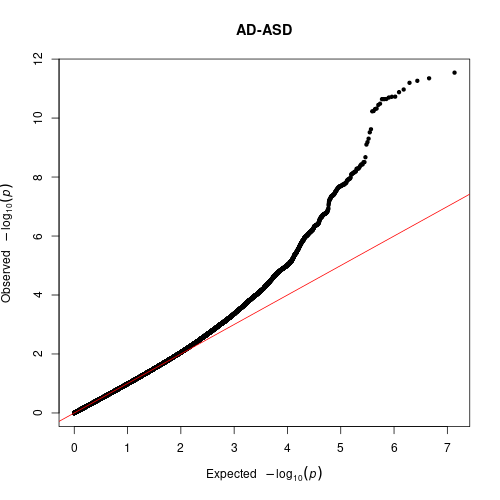

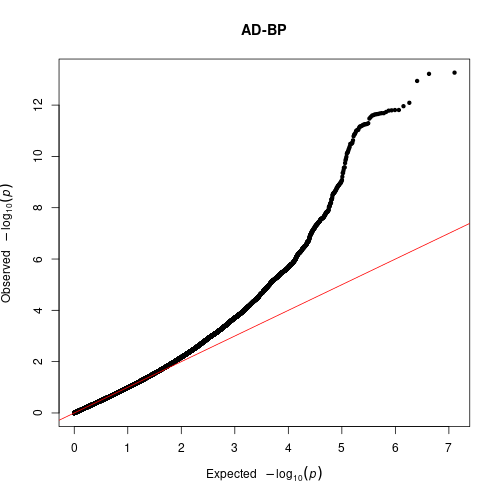

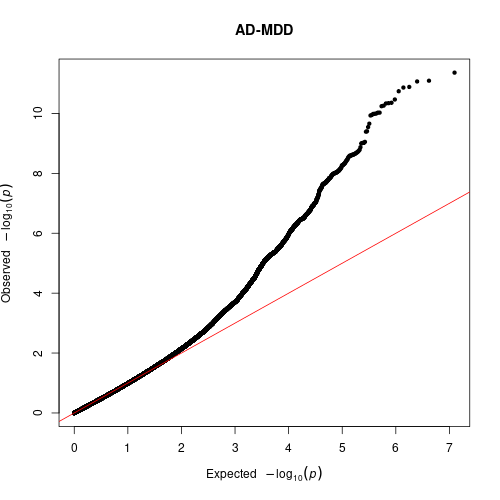

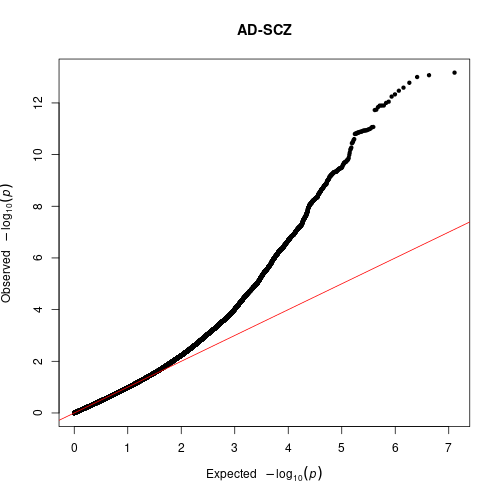

Fig. S2. Locus zoom and locus compare plots for all significant colocalizations in pairwise comparisons.
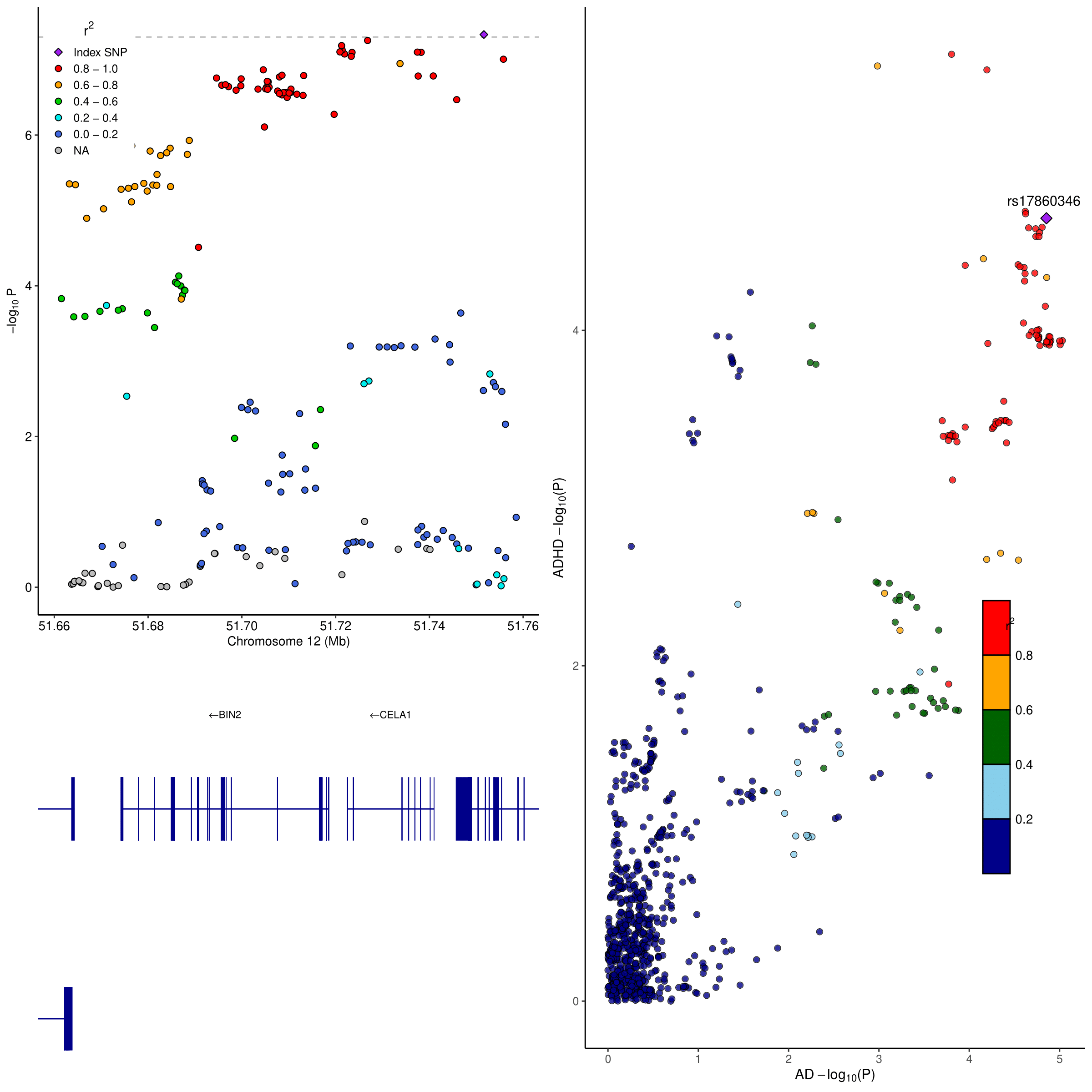

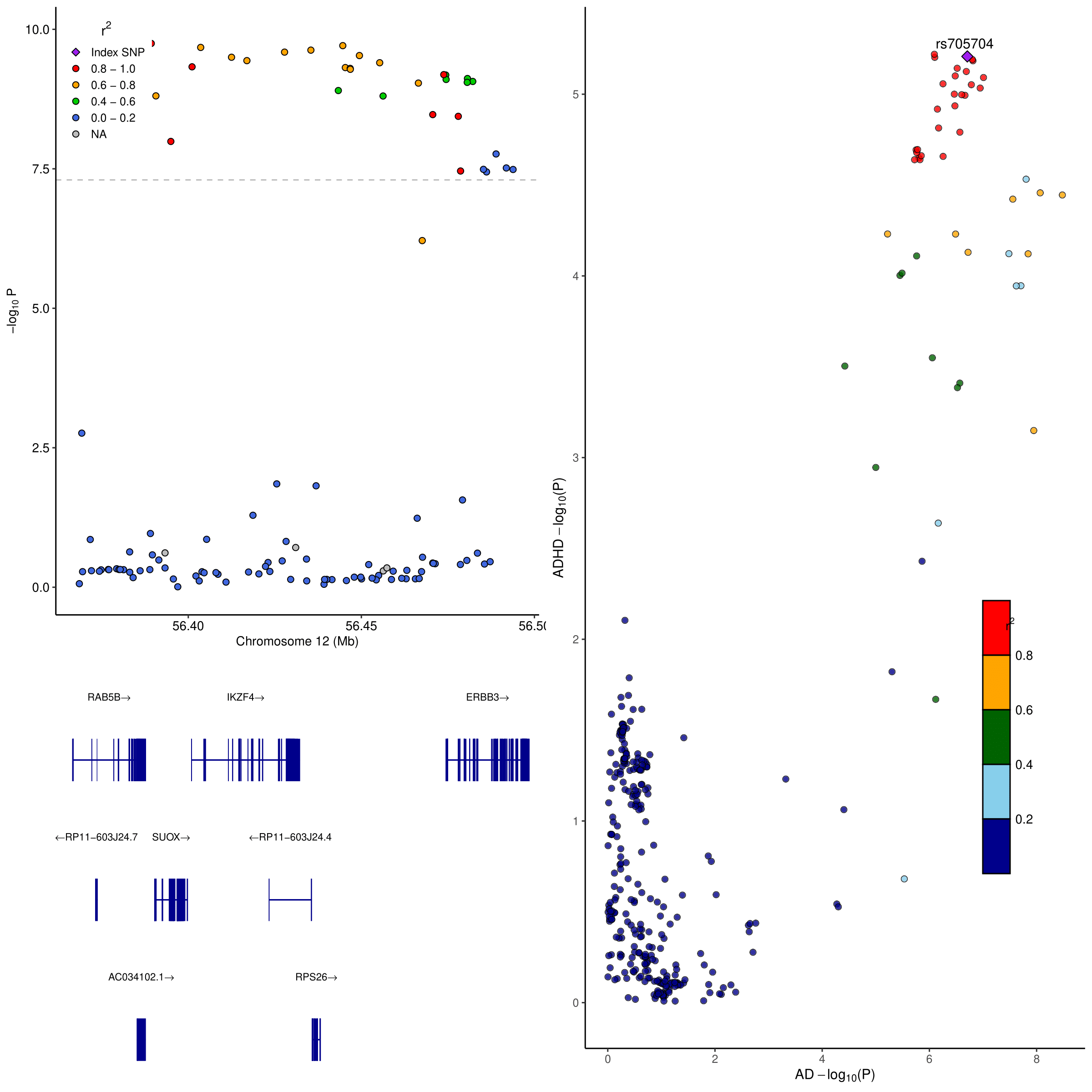

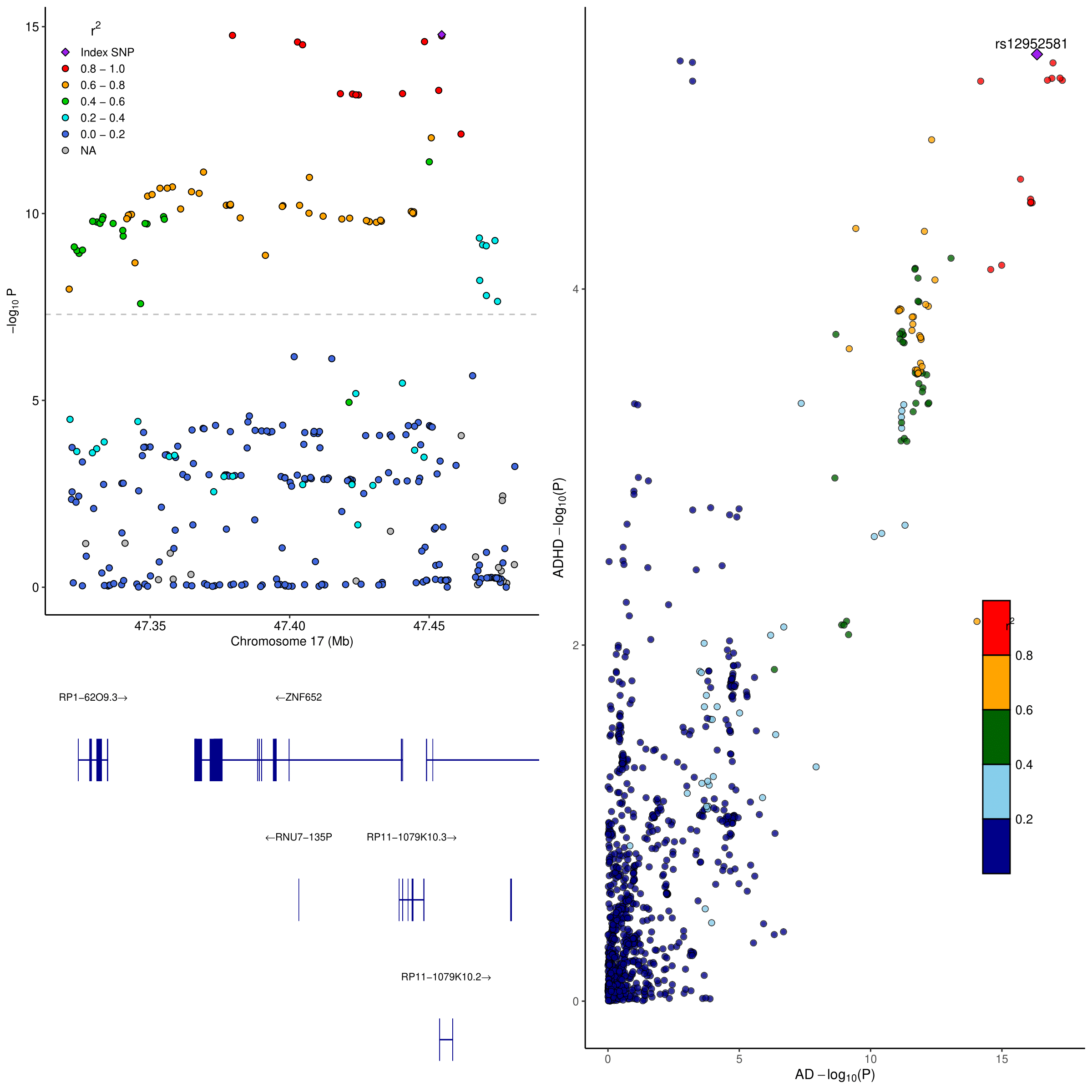

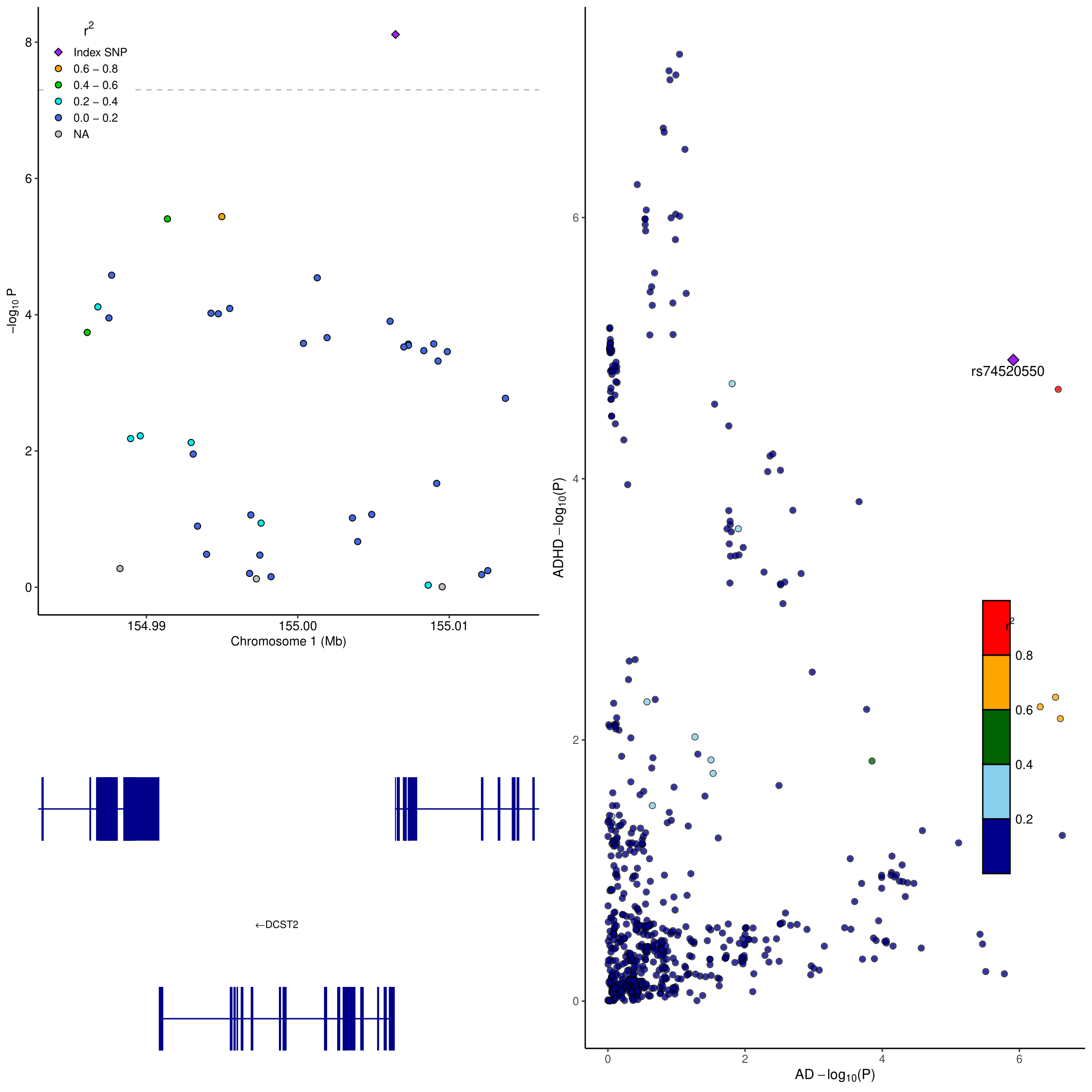

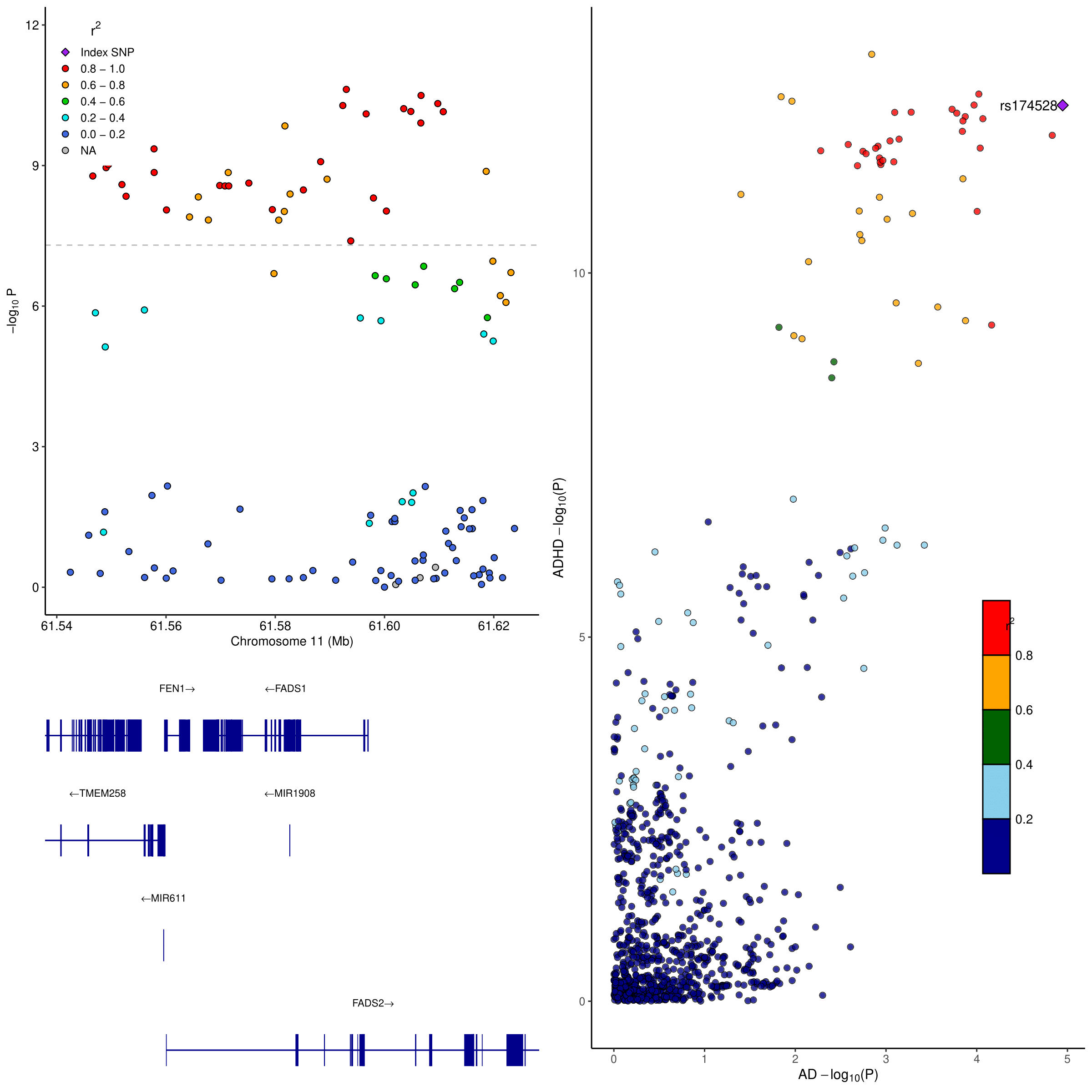

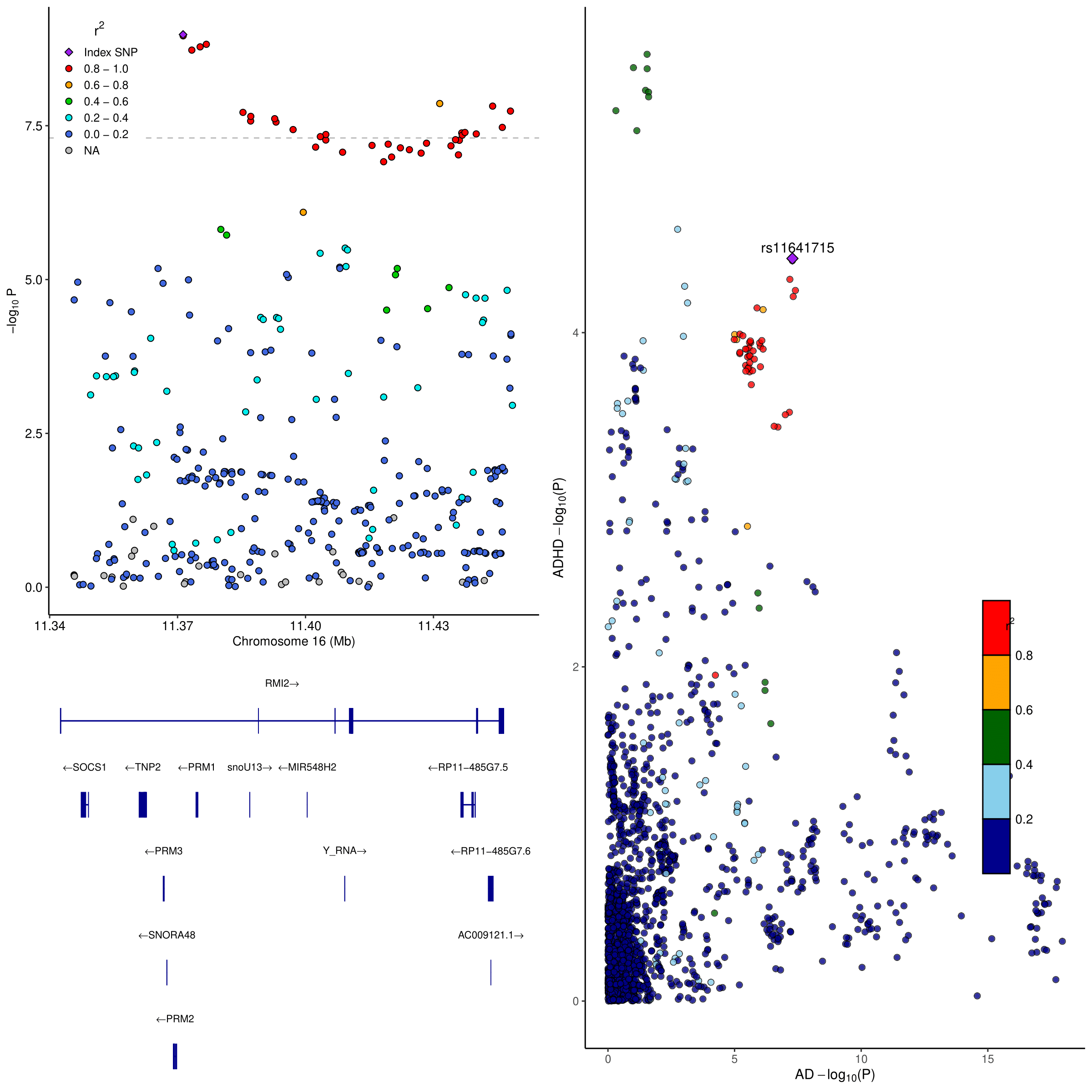

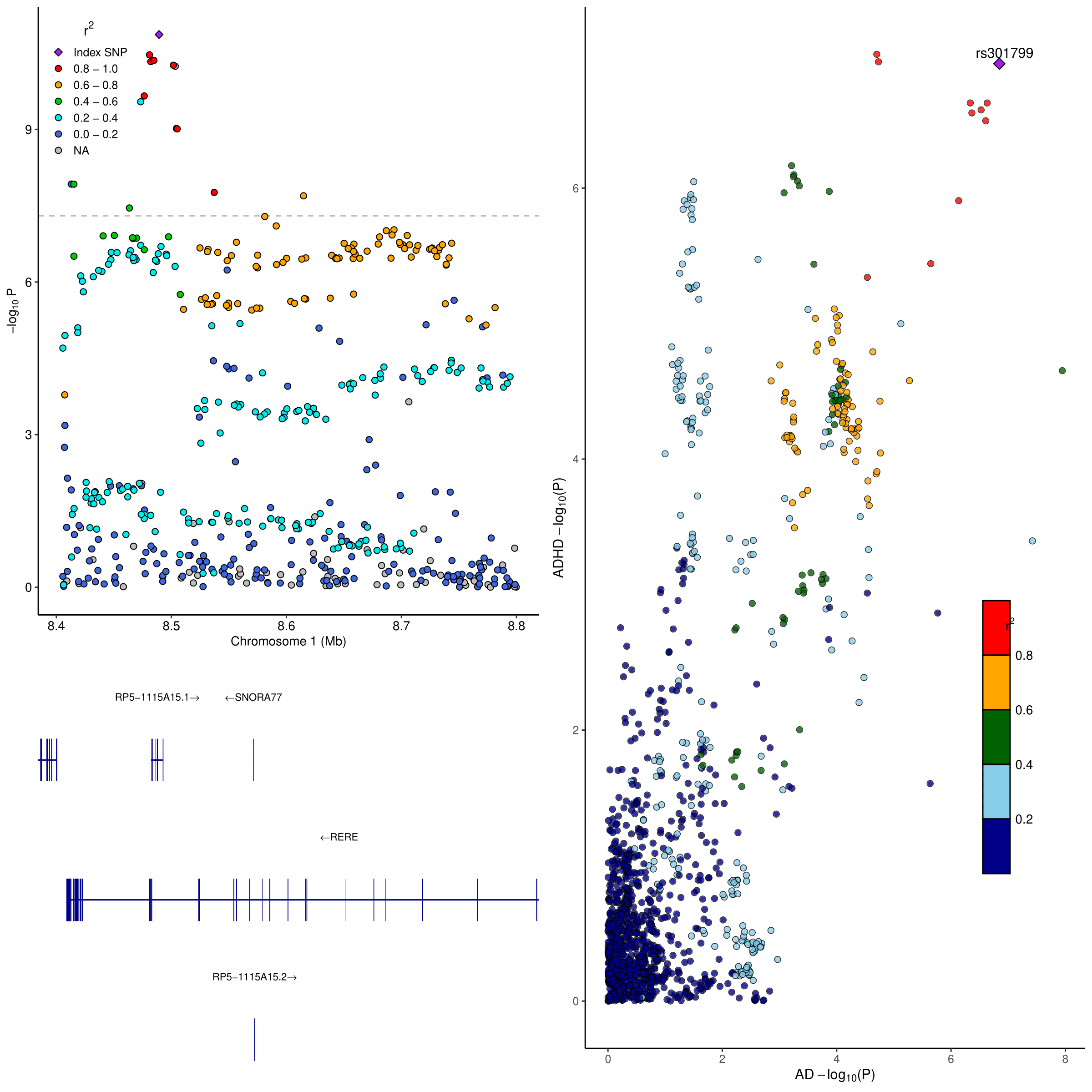

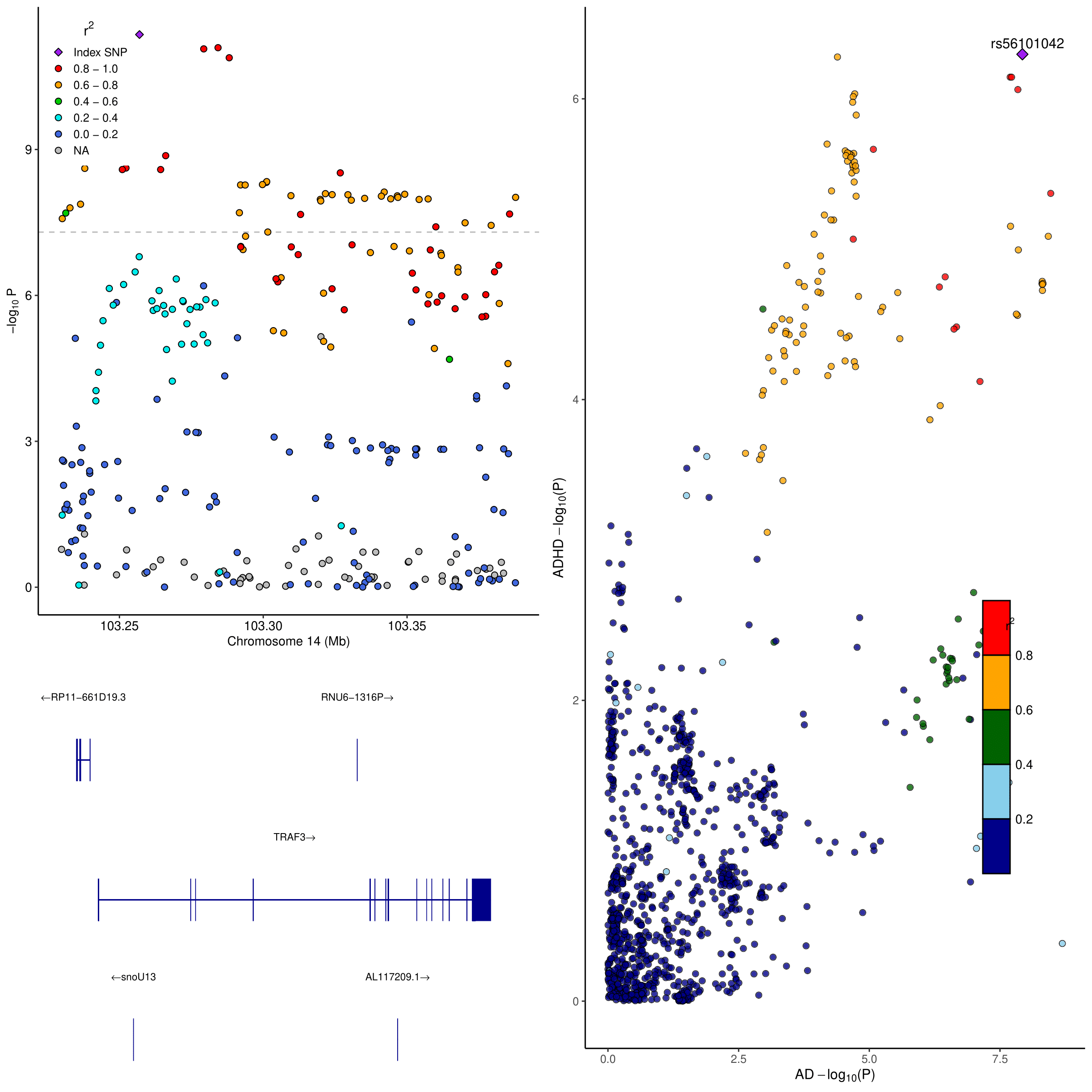

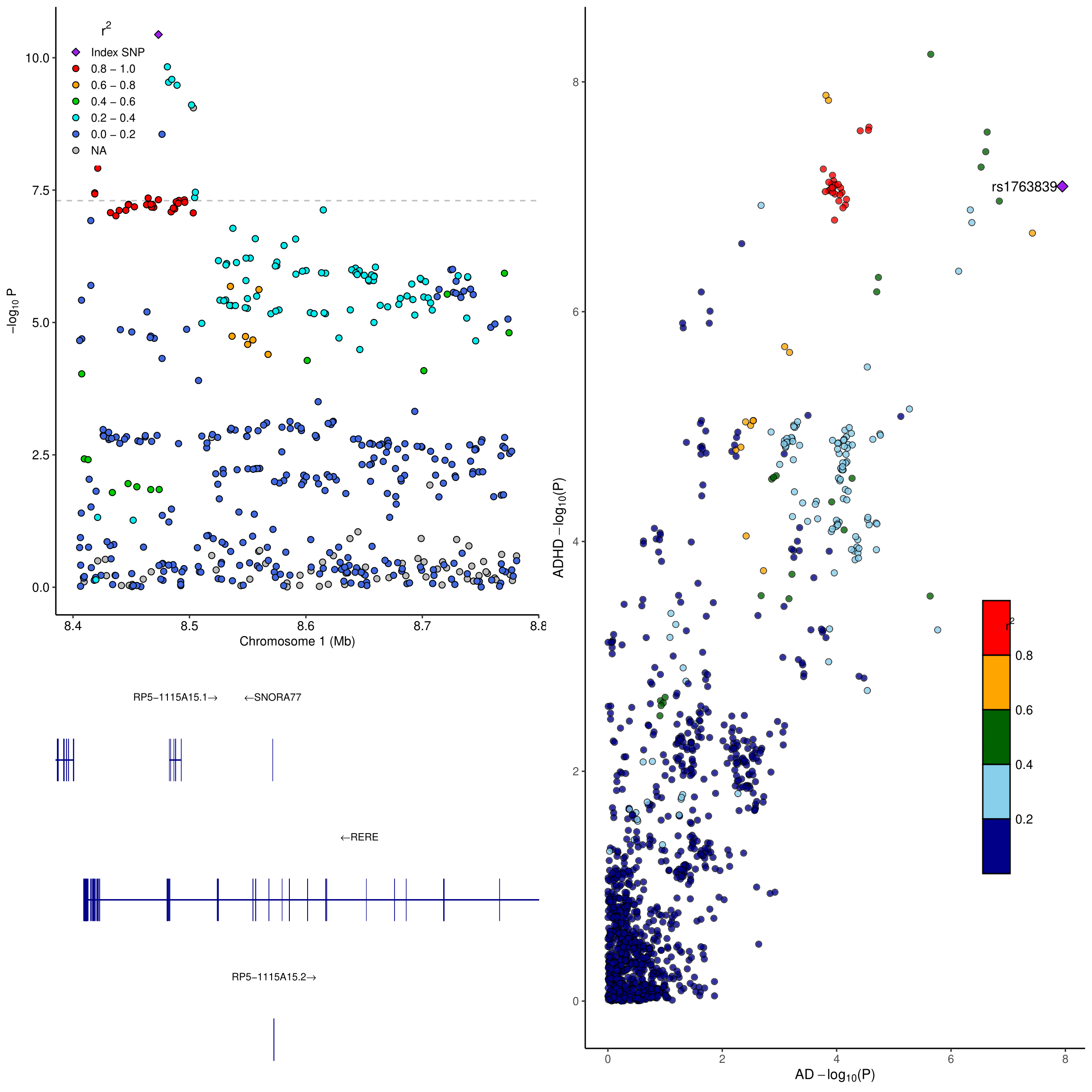

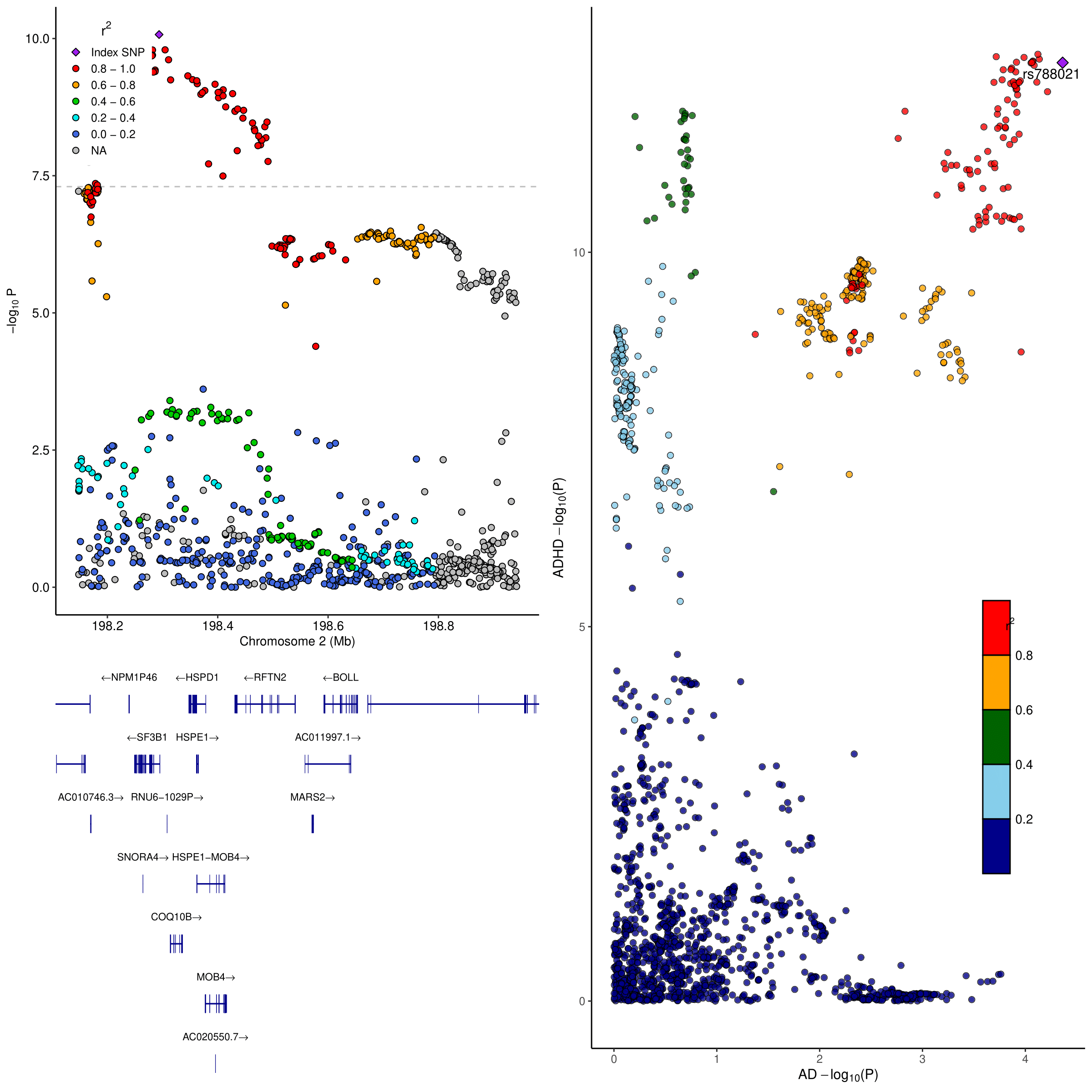

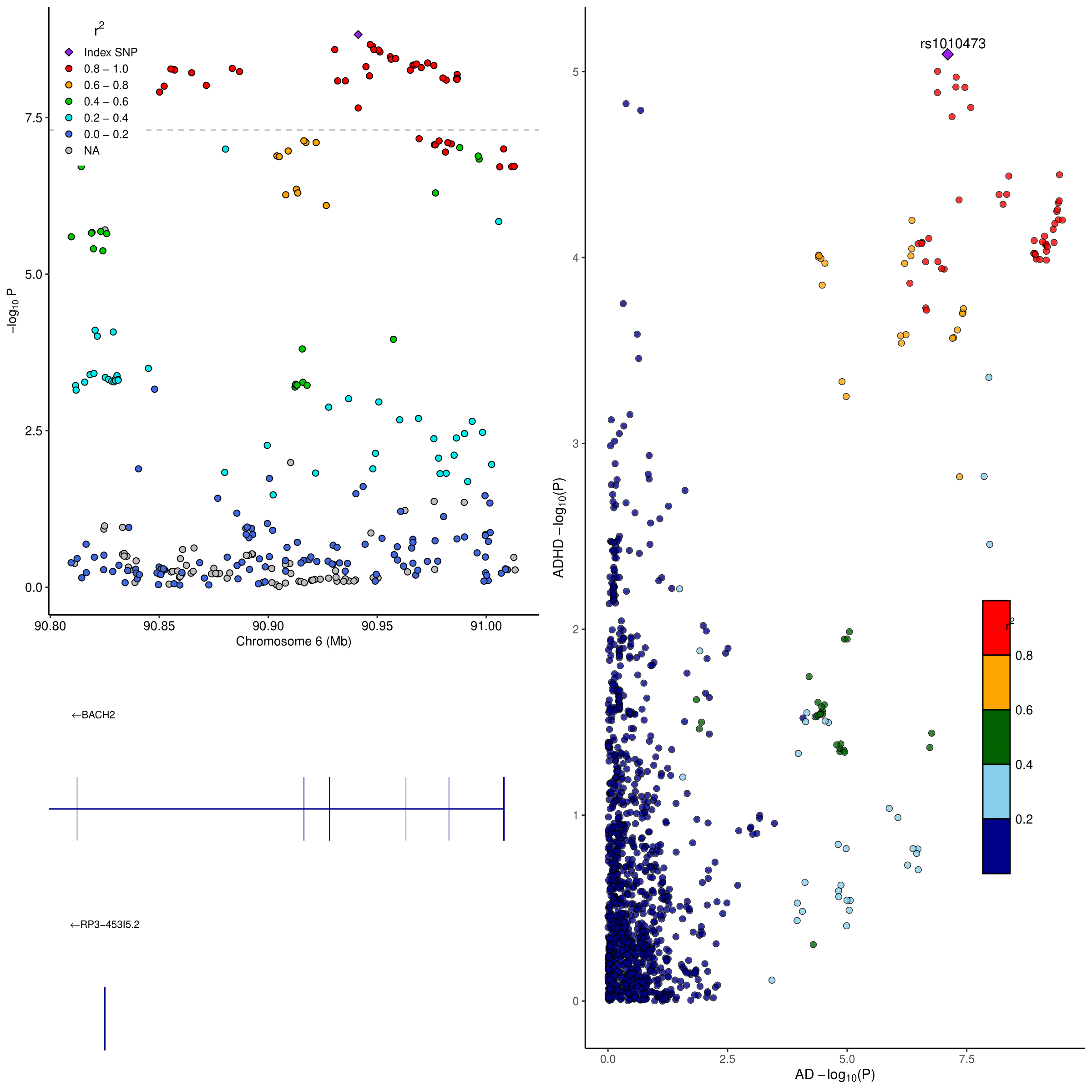

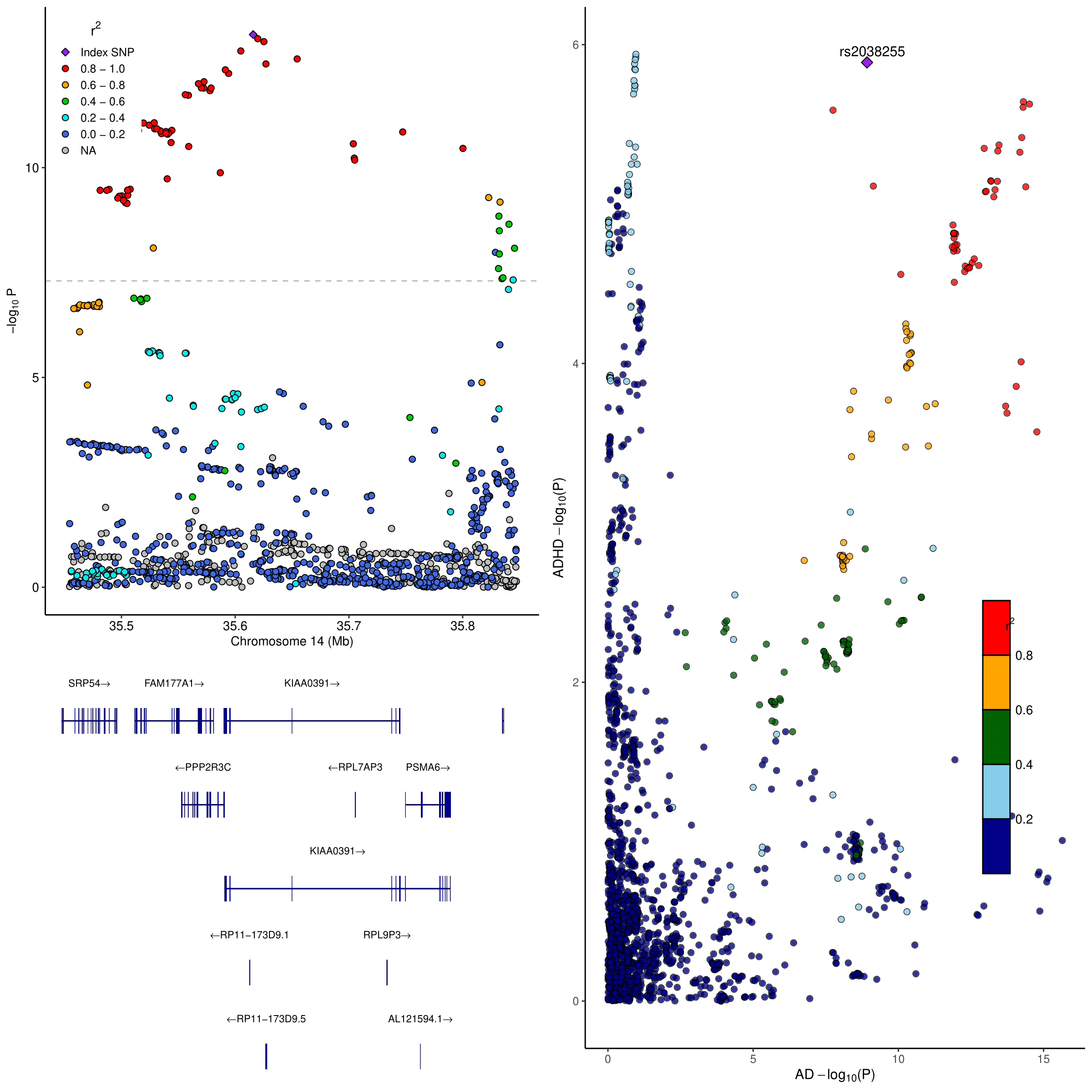

Fig. S3. Pathway crosstalk visualization based on pleiotropic evidence in AD-ADHD pairwise comparison. Parentheses in each node represent the rank in the prioritized gene list.
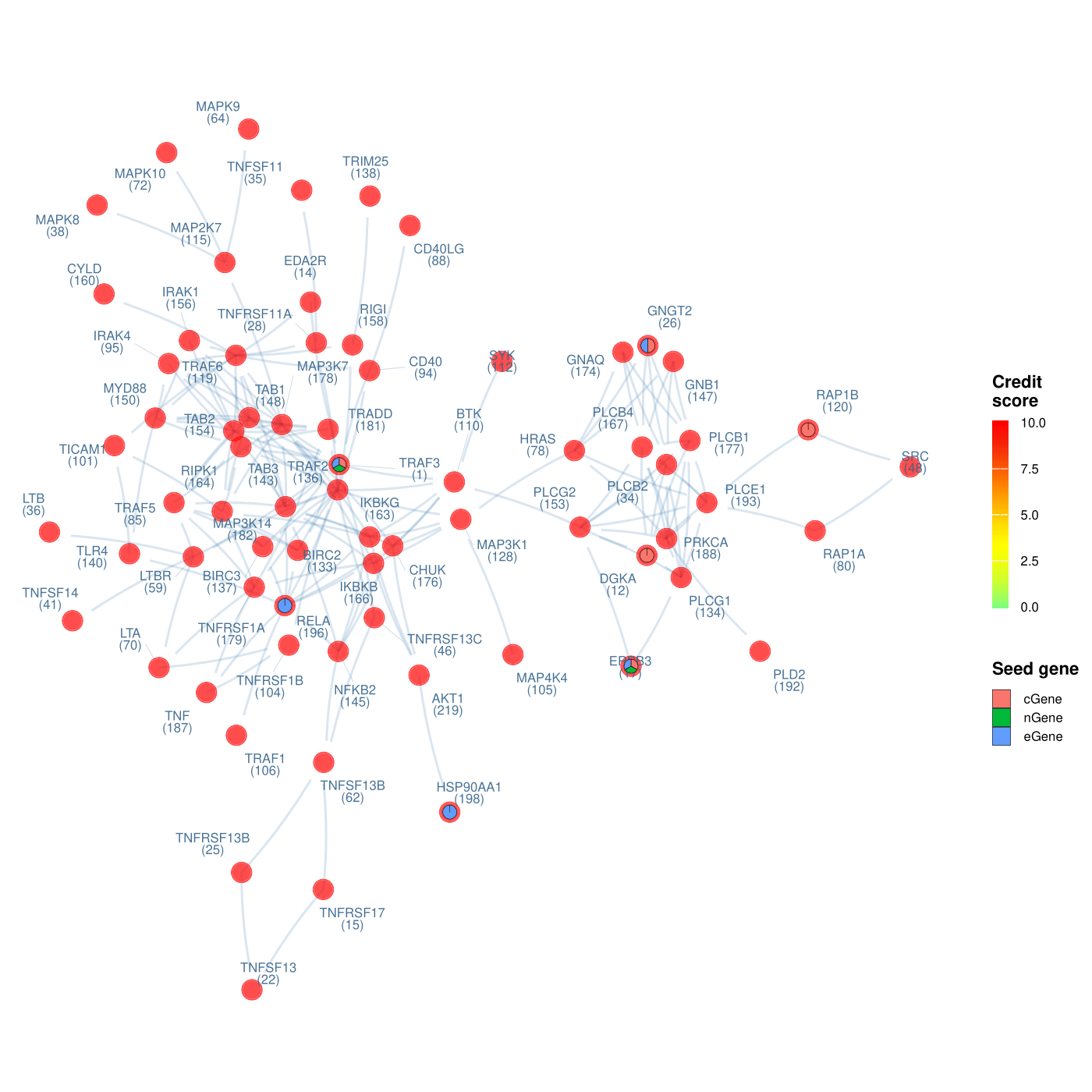

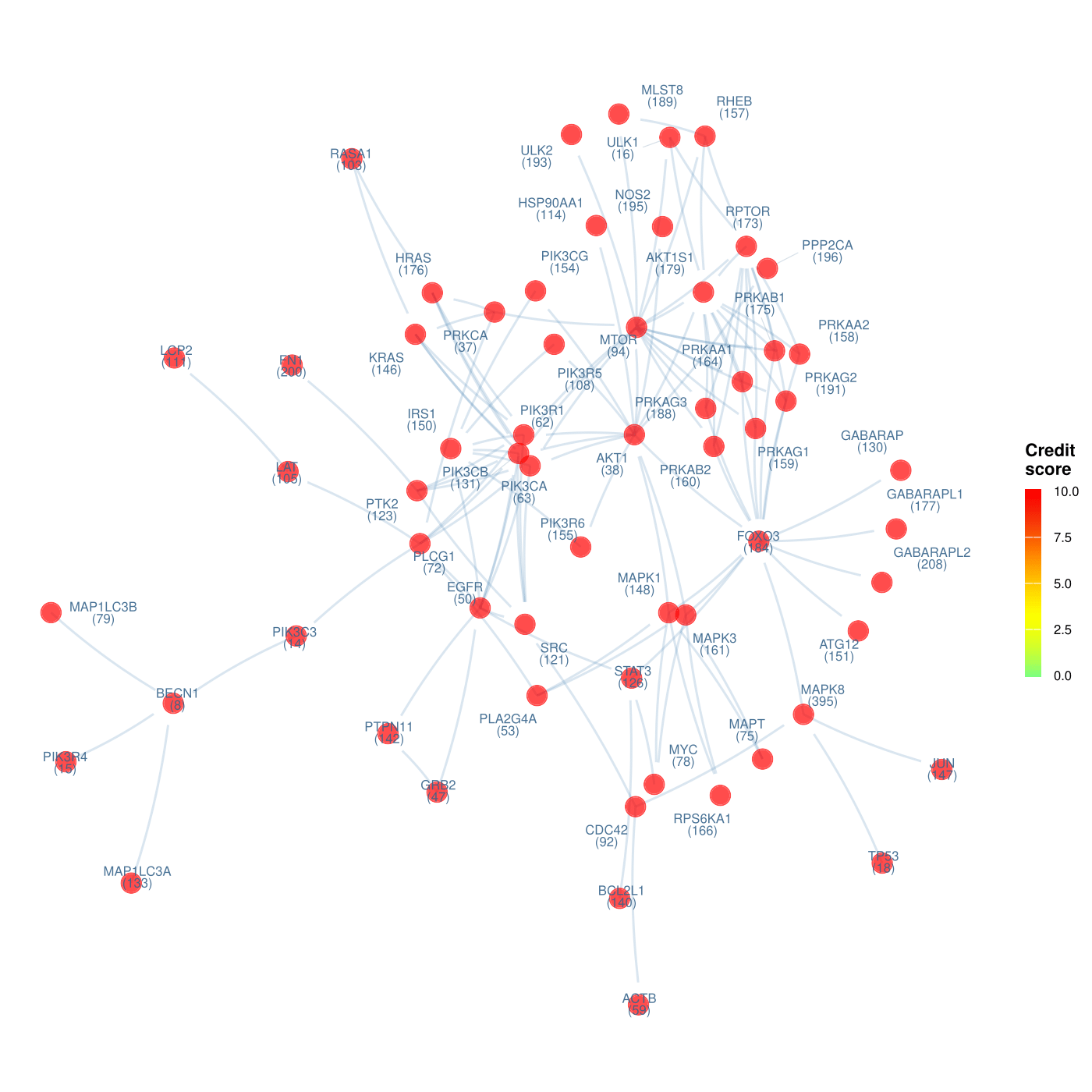
Fig. S4. Pathway crosstalk visualization based on pleiotropic evidence in AD-ASD pairwise comparison. Parentheses in each node represent the rank in the prioritized gene list.

Fig. S5. Pathway crosstalk visualization based on pleiotropic evidence in AD-BP pairwise comparison. Parentheses in each node represent the rank in the prioritized gene list.

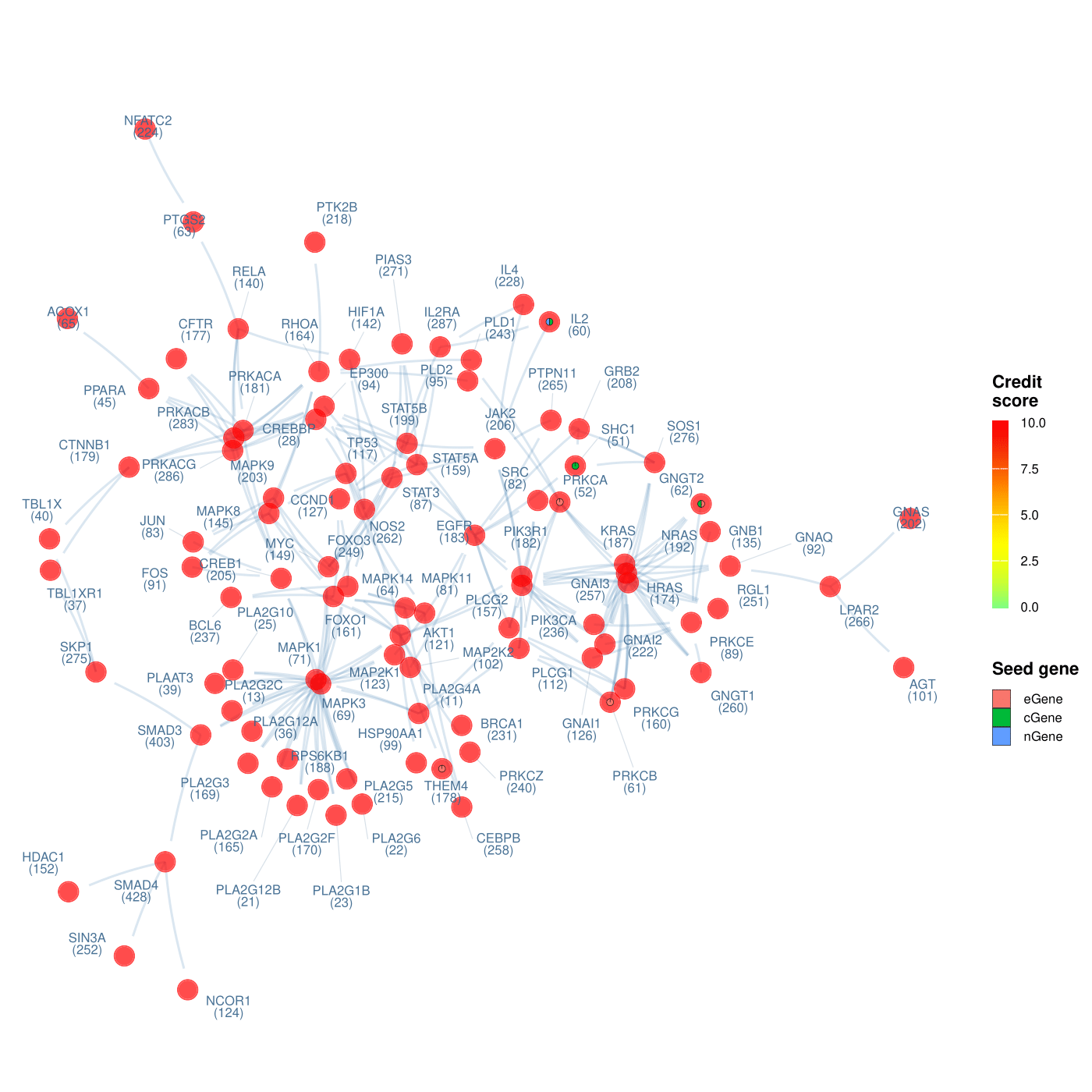

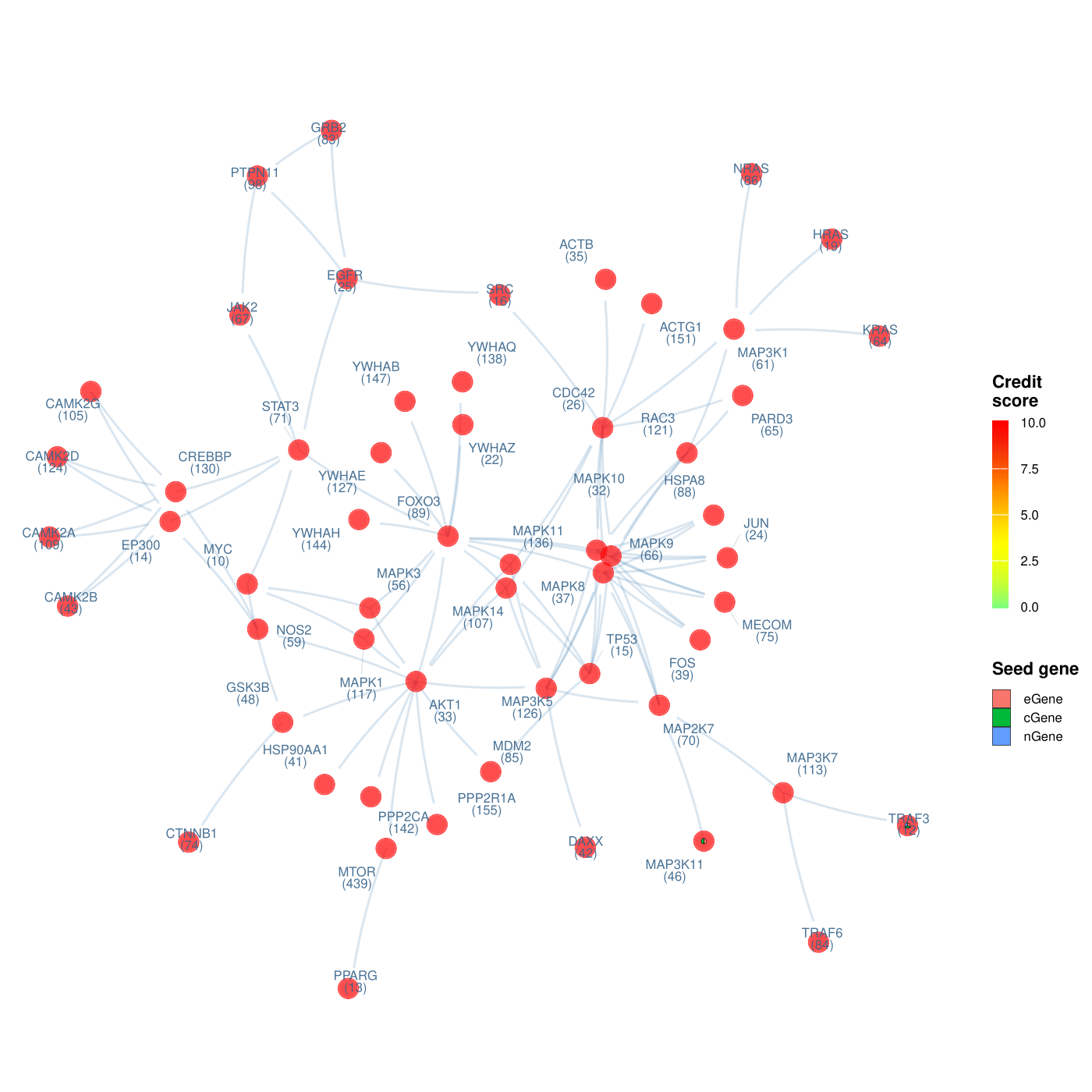
Fig. S6. Pathway crosstalk visualization based on pleiotropic evidence in AD-MDD pairwise comparison. Parentheses in each node represent the rank in the prioritized gene list.

Fig. S7. Pathway crosstalk visualization based on pleiotropic evidence in AD-SCZ pairwise comparison. Parentheses in each node represent the rank in the prioritized gene list.

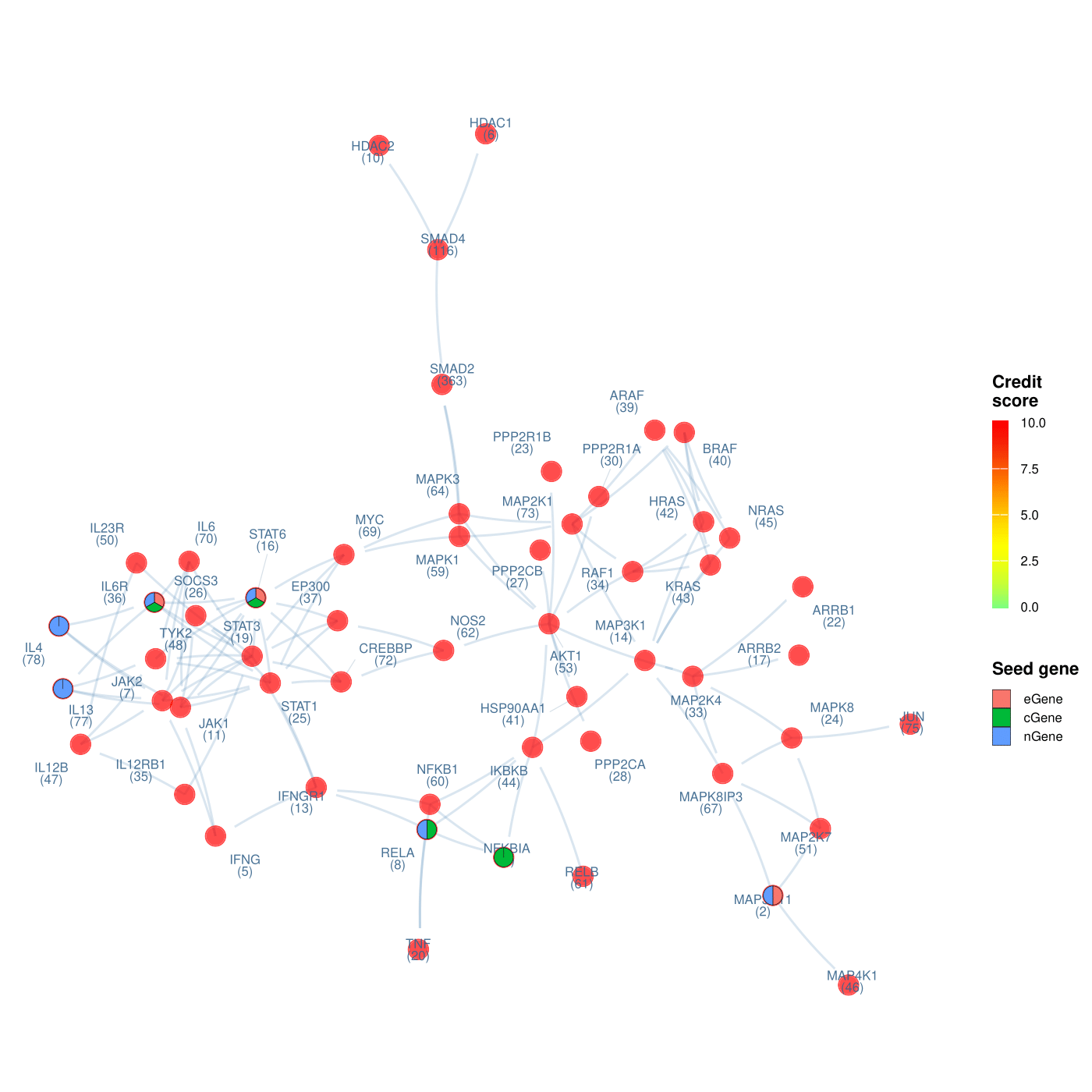

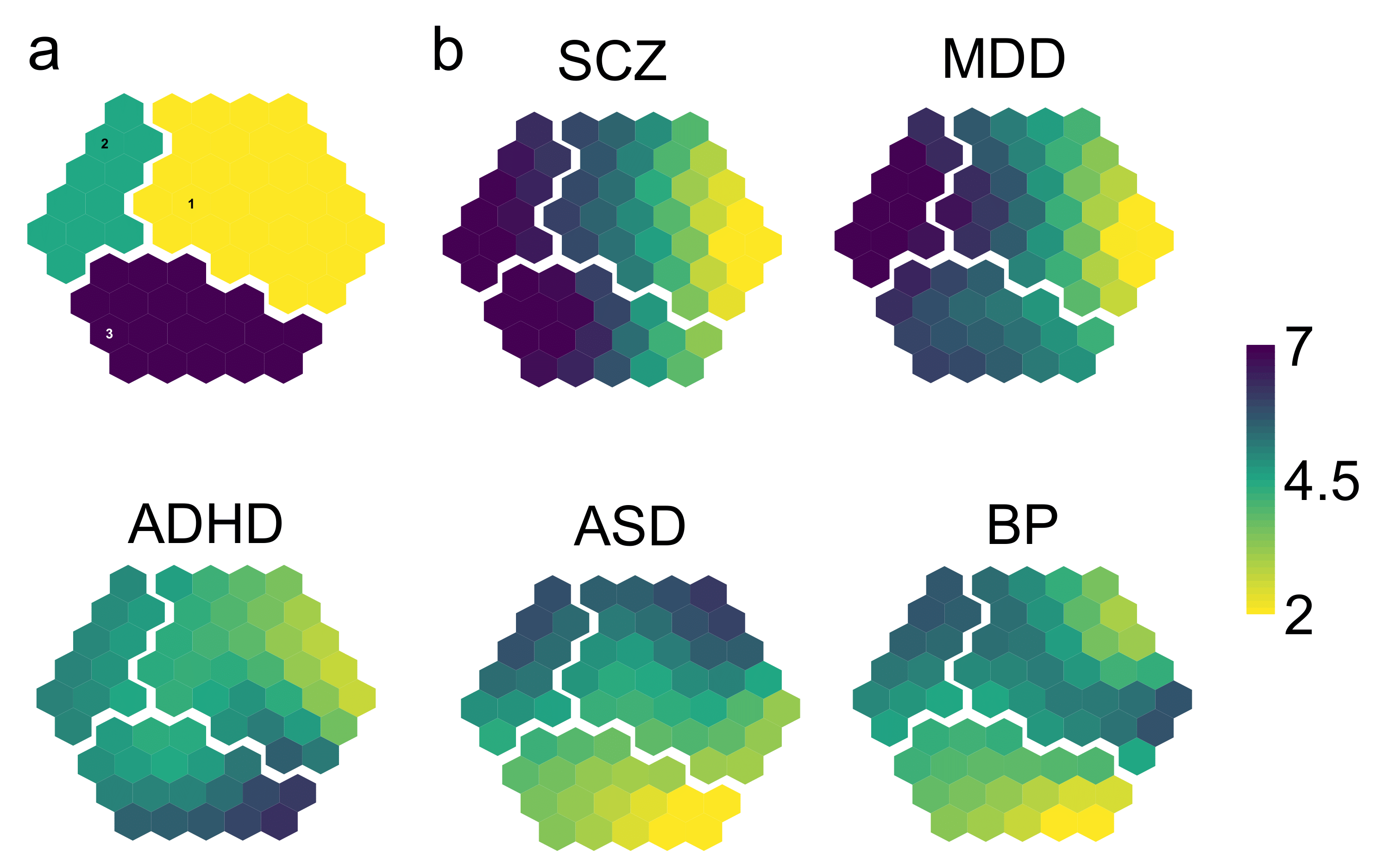
Fig. S8. Pathway crosstalk prioritization map. (a) Illustration of the supra-hexagonal map with mapped all 3 identified clusters. Each cluster is separated by a space. (b) Prioritization map colored by credit score in each pairwise comparison. ADHD, attention deficit hyperactivity disorder; ASD, autism spectrum disorder; BP, bipolar disorder; MDD, major depressive disorder; SCZ, schizophrenia.

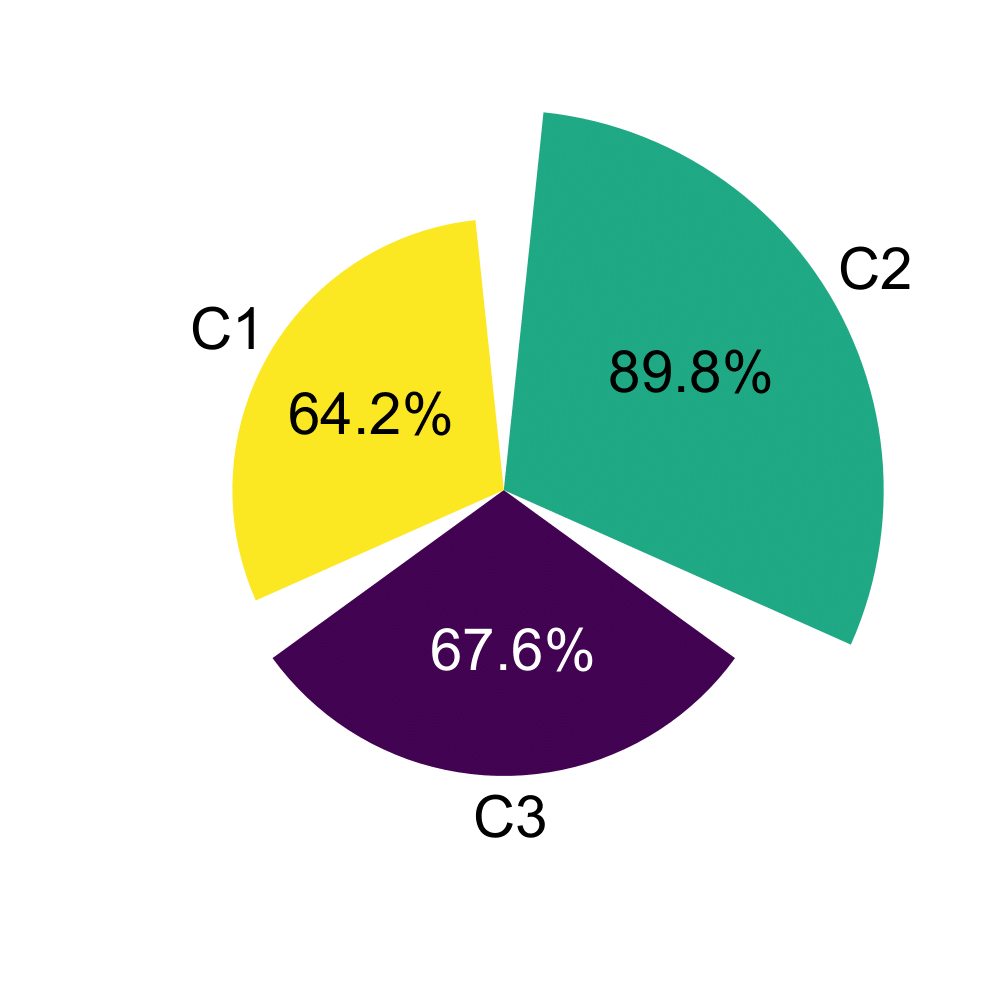
Fig. S9. Proportion of structurally targetable genes in each supra-hexagonal cluster (C1-C3) as identified with the fpocket software.

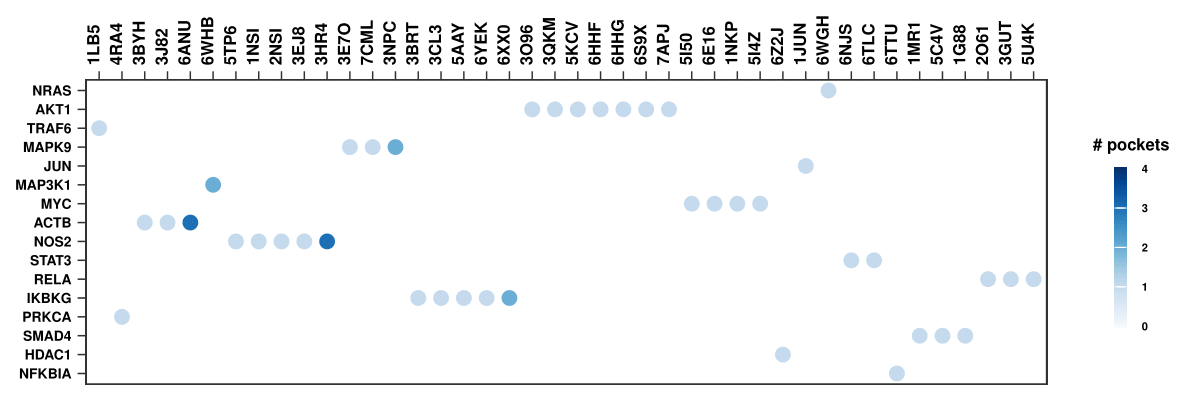
Fig. S10. Structurally targetable proteins in the C2 cluster. The x axis represents structurally targetable domains of each protein with known protein structure through the PDB database in the y axis.

Fig. S11. Mendelian randomization estimates when atopic dermatitis was set as exposure.
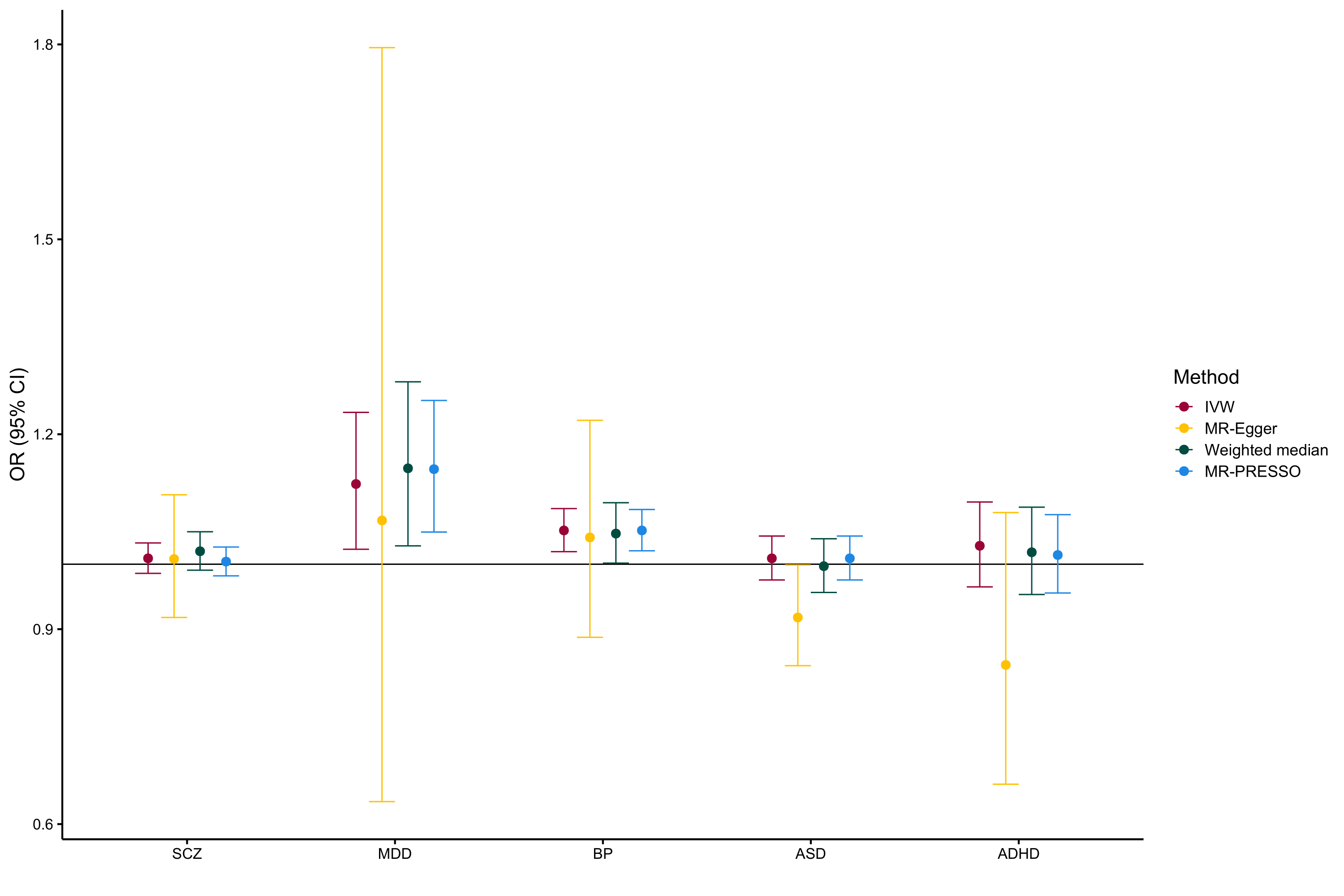

Fig. S12. Mendelian randomization estimates when atopic dermatitis was set as outcome.
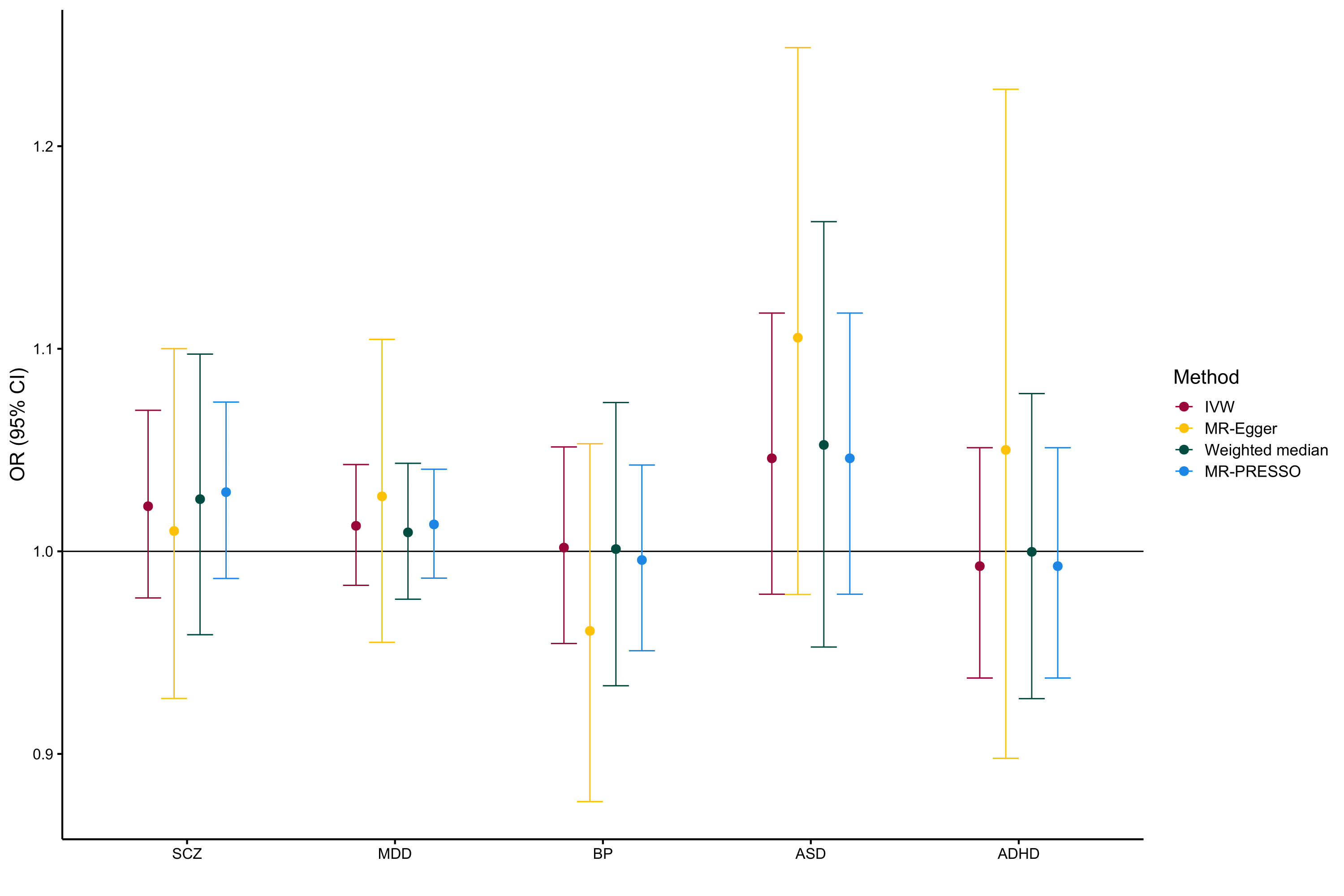
